# No effect of tDCS on fatigue and depression in chronic stroke patients: an exploratory randomized sham-controlled trial combining tDCS with computerized cognitive training

**DOI:** 10.1101/2021.06.22.21258133

**Authors:** Kristine M. Ulrichsen, Knut K. Kolskår, Geneviève Richard, Mads L. Pedersen, Dag Alnæs, Erlend S. Dørum, Anne-Marthe Sanders, Sveinung Tornås, Luigi A. Maglanoc, Andreas Engvig, Hege Ihle-Hansen, Jan E. Nordvik, Lars T. Westlye

## Abstract

Fatigue and emotional distress rank high among self-reported unmet needs in stroke survivors. Currently, few treatment options exist for post stroke fatigue, a condition frequently associated with depression. Non-invasive brain stimulation techniques such as transcranial direct current stimulation (tDCS) have shown promise in alleviating fatigue and depression in other patient groups, but the acceptability and effects for chronic phase stroke survivors are not established. Here, we used a randomized sham-controlled design to evaluate the added effect of tDCS combined with computerized cognitive training to alleviate symptoms of fatigue and depression. 74 patients were enrolled at baseline (mean time since stroke = 26 months) and 54 patients completed the intervention. Self-report measures of fatigue and depression were collected at five consecutive timepoints, spanning a period of two months. While fatigue and depression severity were reduced during the course of the intervention, Bayesian analyses provided evidence for no added effect of tDCS. Less severe symptoms of fatigue and depression were associated with higher improvement rate in select tasks, and study withdrawal was higher in patients with more severe fatigue and younger age. Time-resolved analyses of individual symptoms by a network-approach suggested overall higher centrality of fatigue symptoms (except item 1 and 2) than depression symptoms. In conclusion, the results support the notion of fatigue as a significant stroke sequela with possible implications for treatment adherence and response, but reveal no effect of tDCS on fatigue or depression.

## 1. Introduction

Although recent years have offered considerable improvements in acute stroke care and survival (Walsh, Galvin, Loughnane, Macey, & Horgan, 2015), many stroke survivors experience persistent sequelae (Hankey, Jamrozik, Broadhurst, Forbes, & Anderson, 2002). Described as a “sense of exhaustion, lack of perceived energy or tiredness, distinct from sadness or weakness” (Leegaard, 1983), post stroke fatigue (PSF) is among the most frequently reported (Walsh et al., 2015) and least understood long-term consequences of stroke (De Doncker, Dantzer, Ormstad, & Kuppuswamy, 2018). Fatigue has been shown to increase with time since stroke (Cumming et al., 2018), and a survey of stroke survivors up to five years post stroke identified fatigue, emotional problems and cognitive impairments as the most burdening symptoms (Walsh et al., 2015), indicating a need to target these symptoms in the chronic stroke population.

Generally considered to be a multifactorial condition, PSF is assumed to result from complex interactions between biological, psychological, cognitive, social and behavioral factors (Y. K. Chen et al., 2015; Choi-Kwon & Kim, 2011; Ponchel, Bombois, Bordet, & Hénon, 2015; Wu, Mead, Macleod, & Chalder, 2015; Aarnes, Stubberud, & Lerdal, 2020). The pathogenesis of PSF has not been established (Nguyen et al., 2019), but it is conceivable that individual factors contribute differently during the various stages of recovery. While early fatigue may be associated with biological factors such as stroke severity (K. Chen & Marsh, 2018), lesion characteristics (Tang et al., 2014; Tang et al., 2010) and inflammation (Becker, 2016), fatigue in the chronic phase may be more attributable to behavioral and psychological factors (Wu, Mead, et al., 2015). Cognitive impairments (Passier et al., 2011), including attentional deficits (Radman et al., 2012), reduced processing speed and impaired working memory (Pihlaja, Uimonen, Mustanoja, Tatlisumak, & Poutiainen, 2014) have been reported among PSF patients up to 10 years after stroke (Maaijwee et al., 2015).

The clinical overlap between PSF and post stroke depression (PSD) is substantial (Cumming et al., 2018) - fatigue is both a symptom and a predictor of depression (Douven et al., 2017; van de Port, Kwakkel, Bruin, & Lindeman, 2007), and depressive symptoms in the acute or subacute phase have been associated with increased risk of PSF at 1 to 1.5 years post stroke (Passier et al., 2011; Snaphaan, Van der Werf, & de Leeuw, 2011). While beneficial effects of cognitive behavioral therapy and antidepressants have been reported for PSD (Starkstein & Hayhow, 2019; Wang et al., 2018), a Cochrane meta-analysis concluded that effective interventions for PSF are lacking (Wu, Kutlubaev, et al., 2015), and more information about novel treatments are needed. A recent study on minimally impaired stroke patients reported reduction in fatigue after a single session of anodal transcranial direct current stimulation (tDCS) (De Doncker, Ondobaka, & Kuppuswamy, 2021), a non-invasive brain stimulation technique using low-amplitude direct currents to modulate cortical excitability. tDCS has also been applied to treat PSD (L. Valiengo et al., 2016; L. C. L. Valiengo et al., 2017), and associations between PSD and left dorsolateral prefrontal cortex connectivity or damage (Egorova et al., 2017; Grajny et al., 2016) suggest that neuromodulative methods targeting this region may be particularly effective (Egorova et al., 2017). Yet, the evidence of beneficial effects of tDCS on PSD has been controversial (Bucur & Papagno, 2018), and the mechanisms of potential fatigue-reducing effects remain elusive (De Doncker et al., 2021). Preliminary positive findings should thus be confirmed in larger and controlled studies.

Due to the assumed link between PSF and cognitive impairments, particularly within the domains of attention and processing speed (Johansson & Ronnback, 2014; Ulrichsen et al., 2020), cognitive rehabilitation was recently suggested as a potentially efficient treatment for alleviating fatigue (Aarnes et al., 2020). While significant improvements in fatigue after cognitive training has been reported in patients with multiple sclerosis (MS) (De Giglio et al., 2015), other studies revealed no significant effect on MS fatigue (Pérez-Martín, González-Platas, Eguía-del Río, Croissier-Elías, & Sosa, 2017), and the feasibility of cognitive training for PSF has not been evaluated in chronic stroke patients. Further complicating matters, PSF and PSD may in itself impose considerable barriers to rehabilitation attendance and reduce the probability of positive outcomes (Y. K. Chen et al., 2015; Llorca, Castilla-Guerra, Moreno, Doblado, & Hernández, 2015; Michael, 2002). Investigations of attendance and attrition rates and training gain in relation to fatigue may therefore provide important information for treatment choices.

In sum, effective treatment options for PSF are lacking, but preliminary evidence suggests that tDCS may have a potential to alleviate PSF and PSD. In a trial evaluating the clinical utility of combining cognitive training and tDCS to improve cognitive function post stroke (Kolskaar et al., 2020; Richard et al., 2020), we quantified the effect of real stimulation vs sham on self-reported fatigue and depression using Bayesian mixed effects models. Based on prior literature suggesting beneficial effects of tDCS on fatigue and depression in other conditions, we hypothesized that patients receiving real stimulation would display a larger reduction in symptoms compared to patients receiving sham stimulation. Due to the high comorbidity and symptom overlap between fatigue and depression, we examined the constituents of this relationship in further detail using an exploratory network-approach to map symptom-level centrality and associations at baseline and across time.

## 2. Methods

### 2.1. Sample and study timeline

Figure 1 shows a schematic outline of the study timeline and a flow diagram of recruitment (see Kolskaar et al. (2020) for a detailed description of overall study protocol). Stroke survivors in the chronic phase (> 6 months from stroke onset) were invited to participate. All patients were previously admitted with acute stroke to the Stroke Unit, Oslo University hospital, or the Geriatric Department, Diakonhjemmet Hospital. Exclusion criteria included severe neurological, neurodevelopmental or psychiatric conditions prior to the stroke, MRI contraindications and transient ischemic attack (TIA). All patients suffered mild to moderate strokes (National Institute of Health Stroke Scale (NIHSS; (Meyer & Lyden, 2009) ≤ 7 at hospital discharge), and mean time from stroke onset was 26 months. We included patients with both ischemic and hemorrhagic strokes, but in the current sample, all but one patient suffered ischemic stroke. None of the included patients reported severe linguistic or visual impairments.

**Figure 1.**
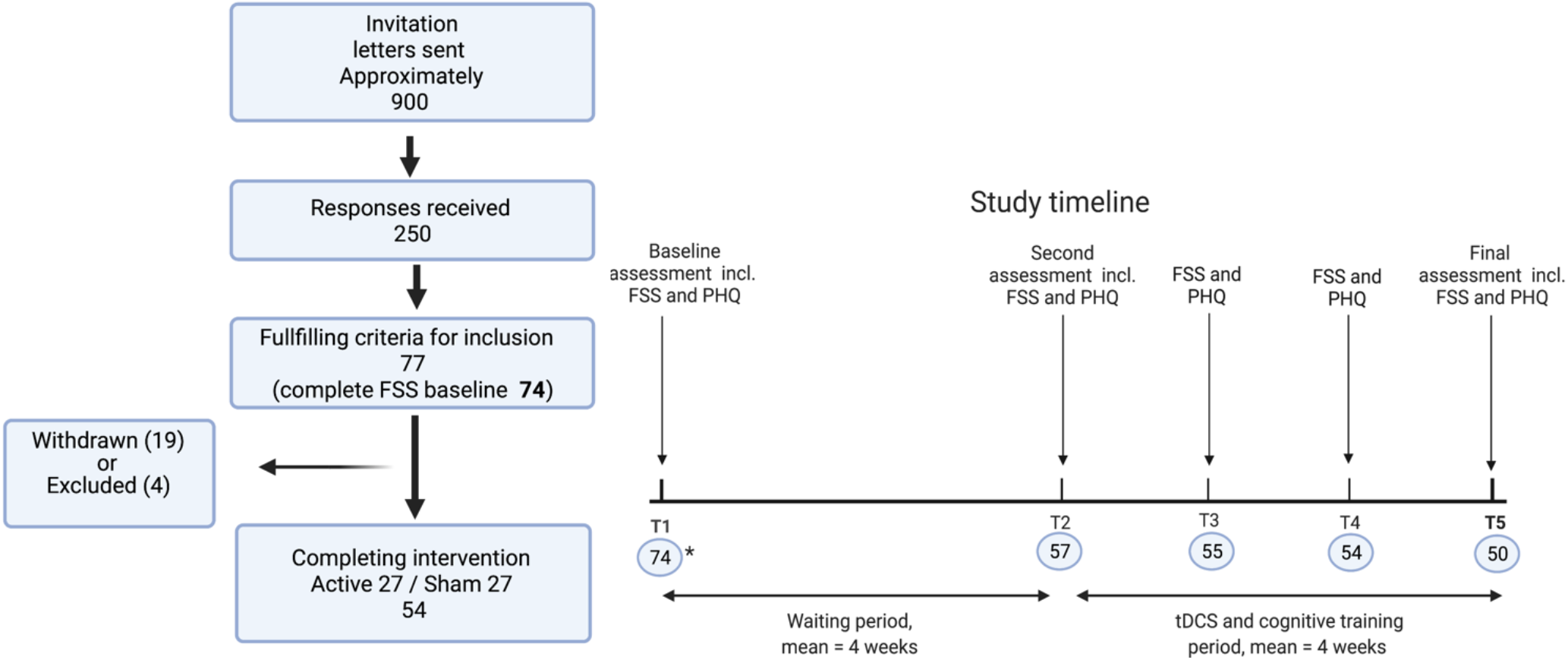
Flow diagram of recruitment (left) and study timeline (right). Number of patients with complete FSS scores is provided in blue circles.

The study was approved by the Regional Committee for Medical and Health Research Ethics South-East Norway (2014/694, 2015/1282). Participants provided written and informed consent prior to enrollment. All participants received a compensation of 500 NOK.

The majority (n = 14) of the 19 patients who withdrew from the study did so prior to the training, while five withdrew during the course of the intervention. None withdrew consent, so baseline data are reported on both completing and withdrawn patients. Three patients were excluded due to medical conditions occurring after inclusion, and one patient were excluded due to problems with fitting the MRI coil.

### 2.2. Cognitive training

Computerized working memory training was done using Cogmed QM (Cogmed Systems AB, Stockholm, Sweden). Details of implementation are described elsewhere (Kolskaar et al., 2020; Richard et al., 2020). Patients completed 17 training sessions over three to four weeks, corresponding to approximately five weekly training sessions. Seven sessions were carried out at Oslo University Hospital, as patients simultaneously received tDCS or sham stimulation (see below) during six of these sessions. The remaining sessions were carried out at home. The Cogmed protocol included 12 different auditory-verbal and visual-spatial exercises, where each session consisted of eight exercises. Level of difficulty adjusts according to initial individual performance, and, to allow for calibration, we did not include the two first sessions for each task in the statistical analyses used to estimate performance gain. Tasks completed less than three times were also discarded. Training gain was calculated for the following eight exercises: Cube, Digits, Grid, Hidden Objects, Rotation, Sort, Twist and 3D-cube.

### 2.3. tDCS protocol

The tDCS protocol is described in detail by (Kolskaar et al., 2020). Briefly, an in-house Matlab script was used to randomize participants into either sham or active condition at study inclusion. A total of six tDCS sessions were administered, at average two times per week, and with a minimum of 48 hours between each session. Active tDCS stimulation was administered at 1 mA, to minimize the risk of adverse effects. Each stimulation lasted for 20 minutes (ramp-up time 120 seconds and fade-out time 30 seconds). The sham stimulation was done by the fade in, short stimulation, fade out approach (Ambrus et al., 2012), with ramp-up followed by 40 seconds of active stimulation before fade-out in accordance with factory settings. Stimulation was delivered a by direct current stimulator (neuroConn DC stimulator plus, Germany), through 5×7 cm rubber pads covered with high-conducting gel (Abralyt HiCl, Falk Minow Services Herrsching, Germany). The anodal electrode was placed over F3 (left DLPFC) and cathodal at O2 (right occipital/cerebellum in the 10-20 system).

### 2.4. Outcome measures

The present study reports on pre-specified exploratory endpoints regarding fatigue and depression in a trial evaluating the clinical utility of combining cognitive training and tDCS to improve cognitive function post stroke (Kolskaar et al., 2020; Richard et al., 2020).

#### 2.4.1 Fatigue and depression self-report measures

Subjective fatigue was measured by the self-report scale Fatigue Severity Scale (FSS) (FSSKrupp, LaRocca, Muir-Nash, & Steinberg, 1989), where impact of fatigue on different areas of daily life is rated from 1 to 7. FSS has demonstrated acceptable psychometric properties (Whitehead, 2009) and is frequently used to assess fatigue in neurological patient populations (Cumming, Packer, Kramer, & English, 2016). FSS scores are usually reported as mean values (lowest mean 1, highest mean 7), where higher scores indicate higher fatigue impact. The cut-off for clinically significant fatigue applied in the literature is either ≥ 4 (Nadarajah & Goh, 2015; Schepers, Visser-Meily, Ketelaar, & Lindeman, 2006) or ≥ 5 (Kjeverud et al., 2020; Lerdal, Wahl, Rustoen, Hanestad, & Moum, 2005; Morsund et al., 2019; Naess, Lunde, Brogger, & Waje-Andreassen, 2012). A cut-off of ≥ 5 has been recommended to prevent overestimation of cases, as a cut-off of ≥ 4 resulted in 42% of healthy controls being identified as fatigued in a large (N=1800) Norwegian sample (Lerdal et al., 2005). However, because both ≥ 4 and ≥ 5 are used in the literature, we here conduct analyses by both values for transparency.

Symptoms of depression were assessed by the depression module of the Patient Health Questionnaire (PHQ-9) (Kroenke, Spitzer, & Williams, 2001). PHQ is a nine-item self-report scale, where items correspond to the criteria of depression as stated in the Diagnostic and Statistical Manual of Mental Disorders (DSM-IV)(American Psychiatric Association, 1994). The respondent indicates degree of symptom load on a scale ranging from 0 (not at all) to 3 (nearly every day), yielding a minimum total score of zero and a maximum score of 27, with ≥10 reflecting moderate depression (Kroenke et al., 2001).

Both fatigue impact and depression were assessed at five time points across an eight-week period (see Figure 1 for timeline). The first assessment was collected approximately four weeks before the second assessment, and the four consecutive assessments were collected on a weekly basis.

#### 2.4.2. Supplementary cognitive measures at baseline

To test for associations between FSS, PHQ and cognition at baseline, the following neuropsychological tests were included: Montreal Cognitive Assessment (MoCA; Nasreddine et al., 2005), the subtests “Vocabulary” and “Matrix Reasoning” from Wechsler Abbreviated Scale of Intelligence, Second Edition (WASI-II; Wechsler, 2011). In addition, the 4-trial version of the Stroop Color Word Interference test (CWIT) was applied to obtain a measure of cognitive speed, inhibition and interference (Delis, Kaplan, & Kramer, 2001). The California Verbal Learning Test (CVLT-II; Delis, 2000) was used as a measure of episodic verbal learning and memory. Here, we included the total number of recalled words across five trials. The Cognitive Failures Questionnaire (CFQ) (Broadbent, Cooper, FitzGerald, & Parkes, 1982) was included as a subjective measure of memory, perception and motor failures.

### 2.5. Statistical Analyses

Statistical analyses were performed using R version 4.0.3 (R core team, 2020).

#### 2.5.1. Effects of tDCS and time on fatigue and depression

To assess effects of tDCS on fatigue and depression, and to quantify evidence in favor of the null and alternative hypothesis, we used Bayesian hypothesis testing. Mixed effect Bayesian regression models were created using the brms package (Bürkner, 2017) in the Stan computational framework (http://mc-stan.org/). We estimated mixed models separately for FSS and PHQ, using FSS or PHQ as dependent variables. Time (1-5), tDCS group (sham, active), tDCS group * time, age, and sex were entered as fixed factors, and participant as random factor. The models were run using 4 chains with 8000 iterations each, in which the first 4000 iterations were discarded as burn-in. Predictors were assigned normal priors with means of 0 and standard deviation of 1. All variables were standardized prior to analysis.

#### 2.5.2. Fatigue and cognitive training – study withdrawal and training gain

Baseline group differences between patients who completed the intervention (n=50), and patients who withdrew from the study (n=19) were first examined by independent samples t-tests. In a follow-up analysis testing for specific effects of PSF (defined as mean ≥ 5/ ≥ 4 on FSS (Lerdal et al., 2005)) on study adherence, we estimated a logistic regression model with completing/withdrawing as outcome variable, and PSF status, PHQ scores, age and sex as predictors.

To quantify individual Cogmed training gain, we followed the approach by Kolskaar et al. (2020). Here, the effect of repeated training was calculated for each subtest by running linear models with task performance as dependent variable and session number as predictor variable for each participant, where the resulting beta-estimates (slopes) reflect the change in performance across time/sessions. Multivariate outlier detection was done by the mvoutliers package in R, using the aq.plot function (Filzmoser & Gschwandtner, 2018). Two of the subtests (“hidden objects” and “digits”) were discarded from further analyses due to a high number of outliers compared to the remaining tests.

We then tested for associations between fatigue and training gain by estimating linear models for each subtest, applying the beta estimate as dependent variable, and FSS score at TP1, age and sex as independent variables. To assess whether potential effects were specific for fatigue, we re-ran the same models with PHQ as predictor variable instead of FSS.

#### 2.5.3. Associations between FSS, PHQ and baseline measures of cognitive performance

To test for associations between fatigue, depression and cognitivive function at baseline, we estimated Bayes factor for linear correlations between FSS and PHQ score, and performance on neuropsychological tests (MoCA, WASI, CVLT, Stroop) as well as subjectively reported cognitive failures (CFQ).

#### 2.5.4. FSS and PHQ associations across time

##### Stability of specific symptoms

To get an estimate of stability and change in individual symptoms across time, we estimated the coefficient of variation (CV) for each FSS and PHQ item across time point 1 to 5, yielding one CV value per item for each person. As FSS and PHQ have different scale properties, the CV values between them cannot be compared directly, but CV estimates provide relevant information about relative stability or change in individual symptoms within each scale.

#### 2.5.5. Network estimations

We used the qgraph package in R (Epskamp, Cramer, Waldorp, Schmittmann, & Borsboom, 2012) to estimate networks based on Spearman’s rank order correlations matrices. We estimated two baseline networks (n = 74), one with sum FSS scores and individual PHQ items to investigate associations between overall fatigue severity and specific depressive symptoms, and one with individual FSS and PHQ items, to visualize item-level associations. The first (sum FSS) network was estimated with regularized partial Spearman correlations via EBICglasso (tuning parameter set to 0.15), while the second (all-item network) was based on full correlations due to the high numbers of parameters relative to sample size and associated stability issues caused by partial correlations. The latter procedure was repeated to estimate five all-items temporal networks with completing patients only, and plotted according to principal component analysis (PCA) dimension loadings, allowing for comparison of network structure across time. PHQ item number 9, suicidal ideation, was excluded from network estimations due to an extremely positively skewed distribution of scores (mean = 0.08). We then estimated one individual-item network based on full correlations for each time point (timepoint 1-5) to investigate item centrality across time. Here, only completing patients (n = 50) was included.

#### 2.5.6. Node centrality and stability

The relative importance of a node (item) in the network can be evaluated by various centrality measures. We estimated strength centrality, which is a stable (Fried, Epskamp, Nesse, Tuerlinckx, & Borsboom, 2016) and commonly examined centrality measure in psychological networks (Malgaroli, Calderon, & Bonanno, 2021), representing the sum of all absolute edge weights directly connected to a given node (Bringmann et al., 2019). To evaluate the relative importance of symptoms across time and networks, we followed the approach by Malgaroli et al. (2021), estimating one network per time point (1-5), and ranked node strength centrality for each time point, before calculating mean across-time centrality ranking for each item. To investigate whether item centrality was associated with symptom severity, we calculated Spearman correlations between mean ranked item centrality and mean item scores.

Stability was assessed using case-dropping bootstrap (nBoots = 1000) by the bootnet package (Epskamp, Borsboom, & Fried, 2018), where network models are estimated on subsets of the data. The correlation stability coefficient (CS) represents the maximum proportion of cases in the sample that can be dropped, maintaining a 95% probability that the correlation between the original centrality scores and the subsets’ centrality scores is minimum 0.70. A commonly applied rule of thumb is that the CV coefficient should not be lower than 0.25, while a coefficient above 0.50 indicates a relatively stable network (Epskamp et al., 2018). Edge accuracy and robustness were tested by bootstrapped estimations of edge confidence intervals and difference test for edges (Epskamp et al., 2018).

## 3. Results

### 3.1. Effect of tDCS and time on fatigue and depression

Figure 2A shows log transformed Bayes Factor evidence plotted for the null hypothesis (intervention has no effect on FSS and/or PHQ) and alternative hypothesis (intervention has effect on FSS and/or PHQ) for all included predictors in the fatigue and depression models. Figure 2B shows the posterior distributions of the coefficients of the standardized predictors for the fatigue model and depression model. Both models provided strong evidence (BF_01_ >10, log-transformed BF_01_ >2) for the null hypothesis of no tDCS treatment effect (no time by group interaction effect on fatigue or depression). Results also provided strong evidence for decreasing symptoms of depression over time (BF_01_ = 0.05, log-transformed BF_01_ < -2), and anecdotal to moderate evidence for reduced fatigue over time (BF_01_= 0.36, log-transformed BF_01_ = -1).

**Figure 2.**
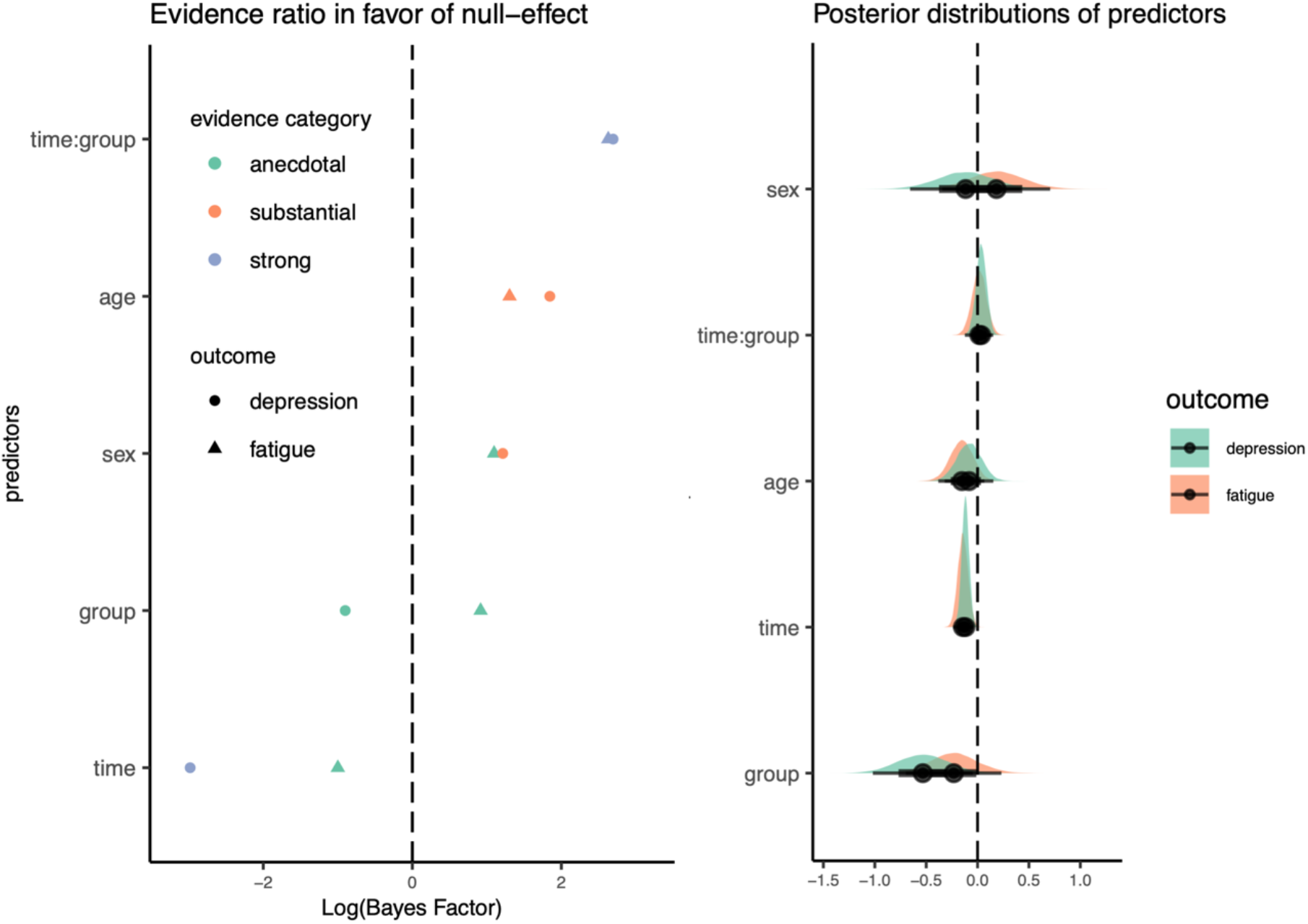
Estimated evidence ratio (A) and Posterior distributions of predictors (B). Log(BF) >0 represent evidence in favor of the null hypothesis and log(BF) < 0 represent evidence in favor of the alternative hypothesis. Posterior distributions of predictors (B) for the fatigue model (red) and depression (green) model.

Figure 3 shows individual FSS and PHQ scores plotted by group for each time point (1-5). FSS mean score for completing patients (N=50) at baseline was 3.5 (SD = 1.5), and 3.0 at TP5 (SD = 1.3). Corresponding sum PHQ values were 4.3 at baseline (SD = 4.2) and 3.5 (SD = 3.3) at TP5.

**Figure 3.**
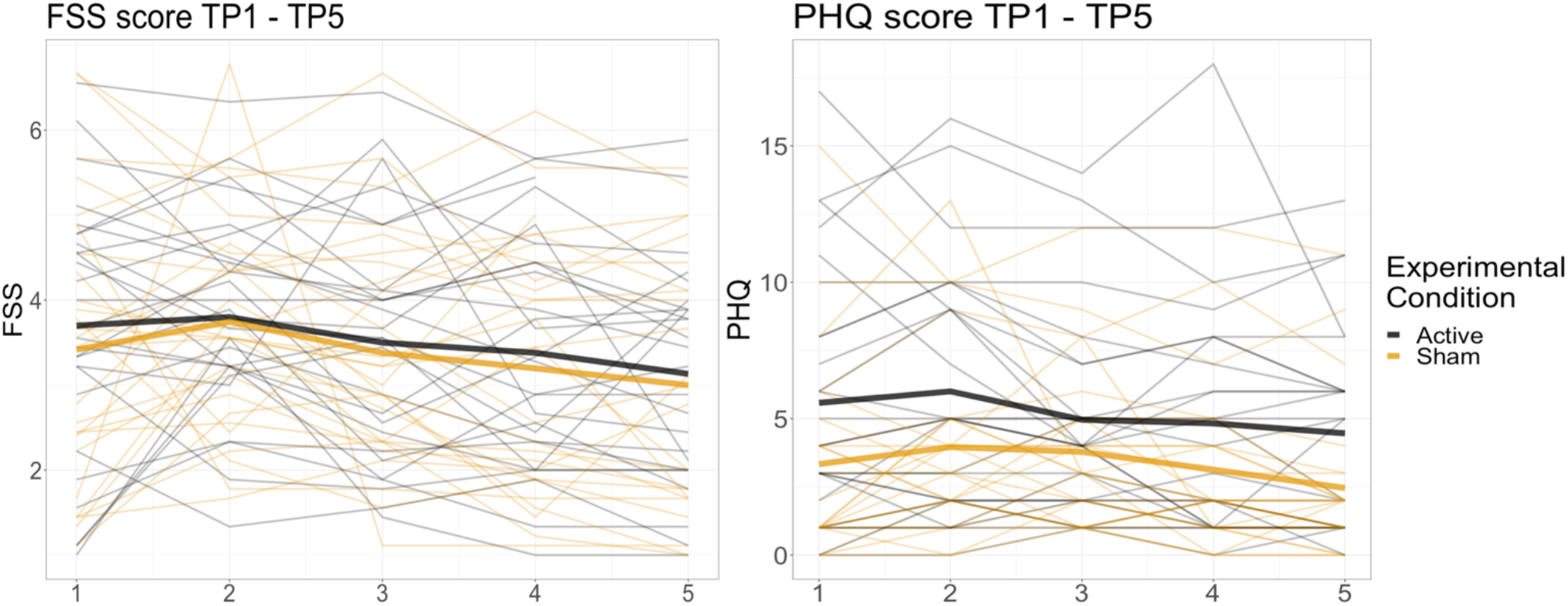
Individual FSS and PHQ scores across time. Scores are grouped by experimental condition (active vs sham).

Follow-up analyses revealed no evidence for a correlation between baseline FSS scores and individual slopes (BF_01_ = 1.33, log-transformed BF_01_ = 0.12), but provided strong evidence for a correlation between baseline PHQ and individual slopes (BF_01_ = 0.001, log-transformed BF_01_ < -2), suggesting that higher baseline PHQ scores were associated with a larger reduction in PHQ symptoms over time.

### 3.2. Fatigue and computerized cognitive training study withdrawal and training gain

Descriptive information on patients who completed the study and patients who withdrew during the study are reported in Table 1. Group differences in continuous variables were tested by independent samples t-tests, and we reported both *t*(*p*) values and Bayes Factors_**10**_, while group differences in sex were tested by Chi-square test of independence.

**Table 1.** Group differences between withdrawn and completing patients.

|  | Withdrawn<br>(n = 19)<br>Mean (SD) | Completing<br>(n = 54)<br>Mean (SD) | Difference withdrawn<br>and completing patients |  |
| --- | --- | --- | --- | --- |
|  |  |  | **t(p)** | **BF<sub>10</sub>** |
| Age | 60.9 (17.1) | 69.1 (7.3) | 2.05 (.051) | **7.7** |
| Sex (chi.sqr) | 10 M/ 9 F | 40 M/ 14F | 2.99 (.083) |  |
| Education | 14.3 (3.9) | 14 (3.7) | 0.38 (.702) | 0.30 |
| NIHSS | 1.3 (1.6) | 1.3 (1.5) | 0.04 (.96) | 0.28 |
| Months since stroke | 29 (7.7) | 25 (9.1) | -1.70 (.093) | 0.70 |
| Lesion volume* | 9239 (15459) | 5978 (9616) | -0.79 (.435) | 0.30 |
| FSS (TP1) | 4.6 (1.5) | 3.5 (1.5) | -2.41 (.022)* | **5.86** |
| PHQ (TP1) | 5.9 (4.2) | 4.3 (4.6) | -1.25 (.190) | 0.69 |
| GAD | 3.2 (2.7) | 2.54 (3.5) | -0.74 (.461) | 0.35 |
| MoCA | 24.7 (4.0) | 25.9 (2.7) | 1.13 (.268) | 0.59 |
| IQ | 110 (16.5) | 110 (16.9) | 0.05 (.954) | 0.30 |

\*One (completing) patient constituted an extreme outlier in terms stroke volume (~8 SDs above the mean) and was removed from the group difference test of lesion volume.

Independent samples t-tests revealed significantly higher fatigue and lower age in patients who withdrew from the study, than in patients who completed the intervention. Follow-up logistic regression analyses including fatigue status (mean score ≥ 5 or ≥ 4 on FSS), PHQ scores, age and sex as predictors for completing/withdrawing from the study, mirrored results from the t-tests. Patients with severe fatigue (mean score ≥ 5 on FSS) were considerably more likely to withdraw from the study compared to patients without fatigue (OR =1.83, 95% CI: 0.27 – 3.52, *p* = .024), but when including patients with moderate fatigue (mean score ≥ 4 on FSS), only age was a significant predictor for withdrawal (OR =0.05, 95% CI: 0.00 – 0.11, *p* = .034).

Linear regression models revealed a significant, negative association between FSS and Cogmed beta slopes in three (uncorrected) or two (corrected) of the six included training tasks (3D Cube, *b* = –0.01, *t*(48) = -2.87, *p* = .006 (FDR-corrected *p* = .021), Grid, *b* = –0.01, *t*(48) = -2.81, *p* = .007 (FDR-corrected *p* = .021), and Sort *b* = –0.01, *t*(48) = -2.09, *p* = .042 (FDR-corrected *p* = .094). However, the effects were small with considerable variation around the regression line, reflected in R^2^ values of .07 and .09. The analyses did not provide evidence for any association between training gain and age, sex or IQ. For illustration purposes, Cogmed slopes are plotted against FSS and PHQ in Supplementary Figure 1. Results from regression models estimated with PHQ as independent variable instead of FSS mirrored the fatigue models, with a significant negative association between PHQ and training slopes identified in two of the same tasks (3D Cube, *b* = –0.004, *t*(48) = -3.92, *p* <.001 (FDR-corrected *p* = .003) and Grid, *b* = – 0.001, *t*(48) = -3.21, *p* = .002 (FDR-corrected *p* = .014). Depression models explained slightly more variance, with R^2^ values of 0.18 and 0.12, respectively).

Formal Bayes Factor model comparison of full model (regression model with either FSS or PHQ included), versus the null model (same model, but without FSS/PHQ score) provided moderate evidence for an association between FSS on learning slopes in 3DCube and Grid, and anecdotal evidence on the sort test. There was no substantial evidence in favor of neither the null (no association with FSS) or the alternative hypothesis (association with FSS) on the remaining tests. The PHQ model revealed a relatively strong association between depressive symptoms on training gain on 3D Cube, Grid and Rotation subtests, indicating smaller gains with higher levels of depression.

Bayes factor estimations for linear correlations (with default priors) provided anecdotal to moderate evidence for no association between fatigue, depression and measures of cognitive function at baseline (MoCA, WASI IQ, CVLT, Stroop, Supplementary Table 2). However, there was substantial evidence for a moderate and strong association between subjectively reported cognitive failures (CFQ) and fatigue and depression, respectively.

### 3.3. FSS and PHQ associations by baseline and across-time networks

Figure 4 depicts individual mean FSS scores at baseline plotted against sum PHQ. Bayes factor estimations for linear correlations provided strong evidence for a positive association between the measures (BF_10_ > 150, mean posterior = 0.68, 97.5 % CI = [0.55-0.79]).

**Figure 4.**
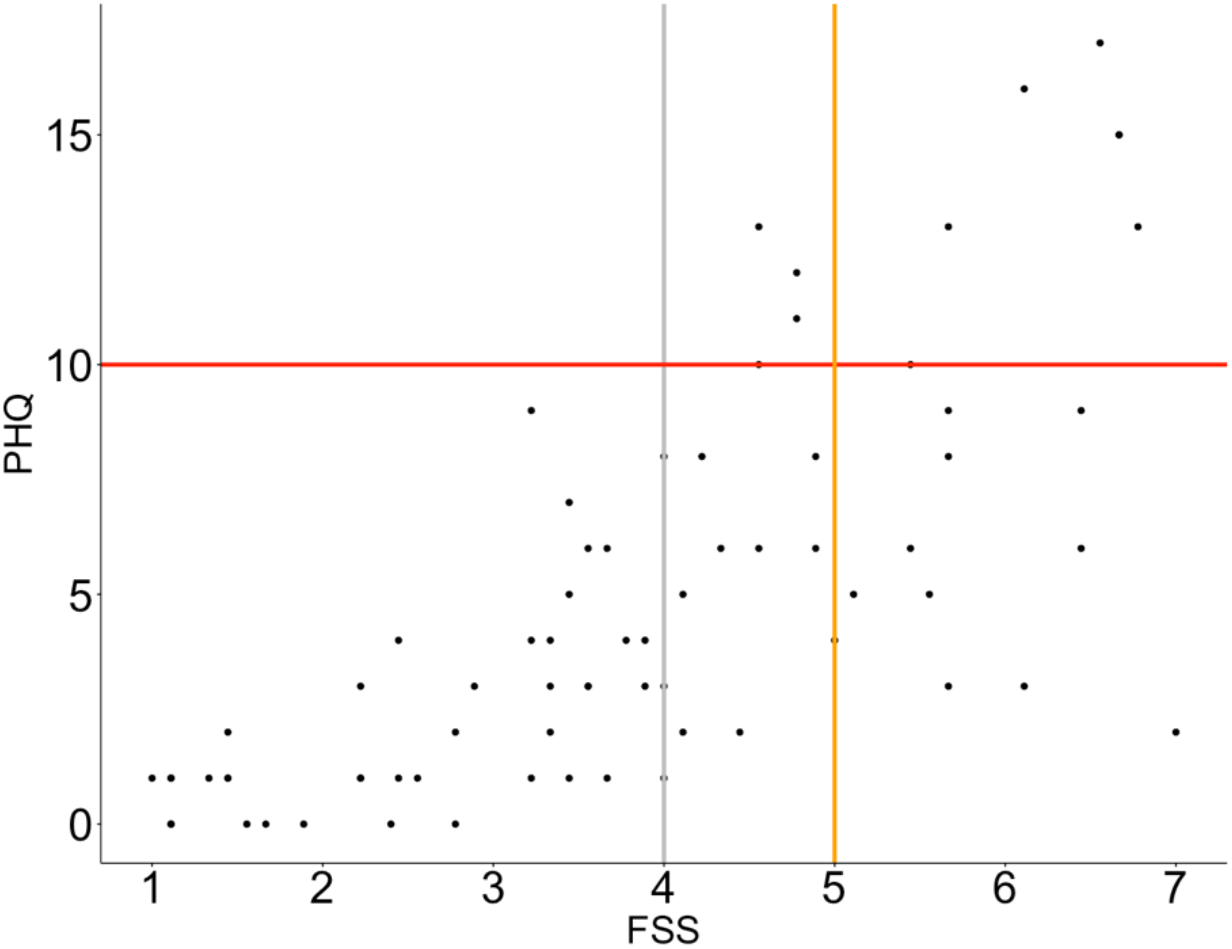
Individual mean scores of FSS (x axis) and PHQ sum scores (y axis) plotted for all patients (n = 74). Vertical lines (grey and orange) mark commonly used cut off values for clinical fatigue, while horizontal red line marks cut off value for depression.

All patients scoring above clinical cut off for moderate depression on PHQ, also experienced moderate or severe fatigue. The association was not reciprocal, in that several patients reported high FSS scores, without displaying clinical levels of depression. Supplementary Table 1 shows the individual items in FSS and PHQ, along with the mean aggregated scores (TP1 – TP5), standard deviations and coefficient of variation (CV) values, indicating degree of variability across time points.

Figure 5 shows baseline networks estimated with FSS sum scores and individual PHQ items (left), and all FSS and PHQ items (right). The sum FSS graph indicated that PHQ items reflecting tiredness, lack of energy and trouble concentrating showed strongest associations with overall fatigue. However, only one edge “sum FSS – PHQ 4 (tiredness/lack of energy)” was identified as significantly different than the majority of other network edges by bootstrapped difference test for edge-weights (Supplementary Figure 2). Item strength centrality (CS-coefficient) for network 1 (sum FSS and PHQ scores) at baseline was estimated to 0.28 by case-dropping bootstrap sampling, meaning that up to 28% of the sample could be dropped while retaining a correlation of 0.70 with the original sample strength centrality (95% CI). Corresponding CS-coefficient for baseline network based on full correlations between all items (right) was estimated to 0.51. Bootstrapped difference test for edge-weights revealed that most edges were not significantly different from the majority of other edges (Supplementary Figure 3), with the strongest edge being FSS 8 (“Fatigue is among most disabling symptoms”) and FSS 9 (“Fatigue interferes with my work, family or social life”). Bootstrapped confidence intervals (CIs; Supplementary Figure 4) showed a substantial overlap between edge-weights, indicating that order of edges should be interpreted with caution (Epskamp et al., 2018).

**Figure 5.**
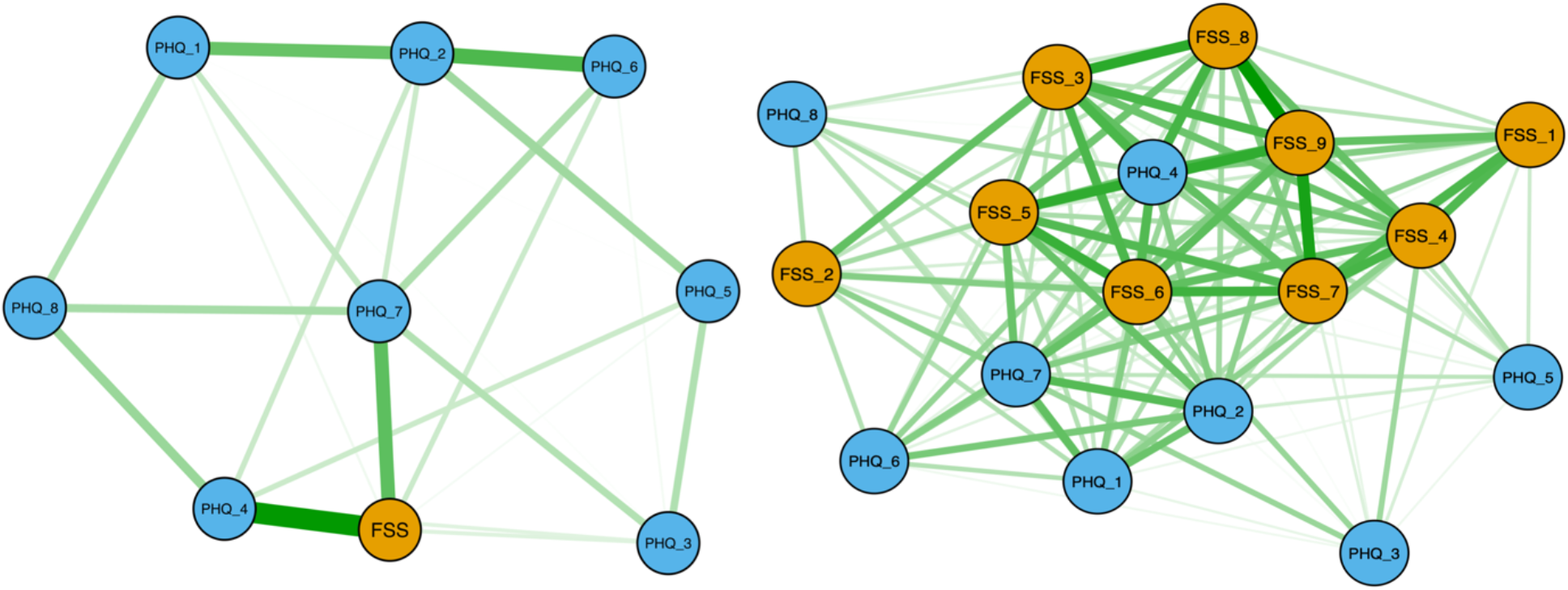
Associations between baseline FSS and PHQ. Network visualization of Spearman partial correlations with EBICglasso regularization (tuning parameter = 0.15), between FSS sum score (FSS_1) and PHQ items for all patients (n = 74) at baseline (left). Network visualization of full Spearman correlations between all FSS and PHQ items at baseline (right). Green edges signify positive correlations, while red edges (none present) represent negative correlations (Epskamp et al., 2012). The thickness of the lines indicates the strength of the association.

Temporal networks based on full correlations are shown in Supplementary Figure 5, and corresponding plots for bootstrapped difference test for node strength in Supplementary Figure 6. Nodes are placed according to loadings on unrotated PCA dimensions. While this approach should be considered exploratory due to the high number of items relative to sample size, the temporal network graphs show that associations between symptoms/network structure vary across times of measurement, and suggest that fatigue- and depression items tend to cluster according to their’ respective scales.

Figure 6 shows estimated standardized strength centrality (left) and ranked centrality (right) for individual items at each time of measurement, suggesting acceptable consistency across time for most items, except PHQ item 7 (concentration problems), 5 (appetite), 4 (tired/little energy) and 6 (feeling bad about yourself). Evidence for an association between overall symptom severity (calculated as mean item score across time 1-5) and overall item centrality (estimated as mean ranked centrality across time) was moderate (BF_10_ = 3.1, mean posterior = -0.40, 97.5 % CI = [-0.71 – 0.03]) with wide credible intervals, indicating that higher symptom severity is associated with increased symptom centrality, but that the strength of the relationship is uncertain.

**Figure 6.**
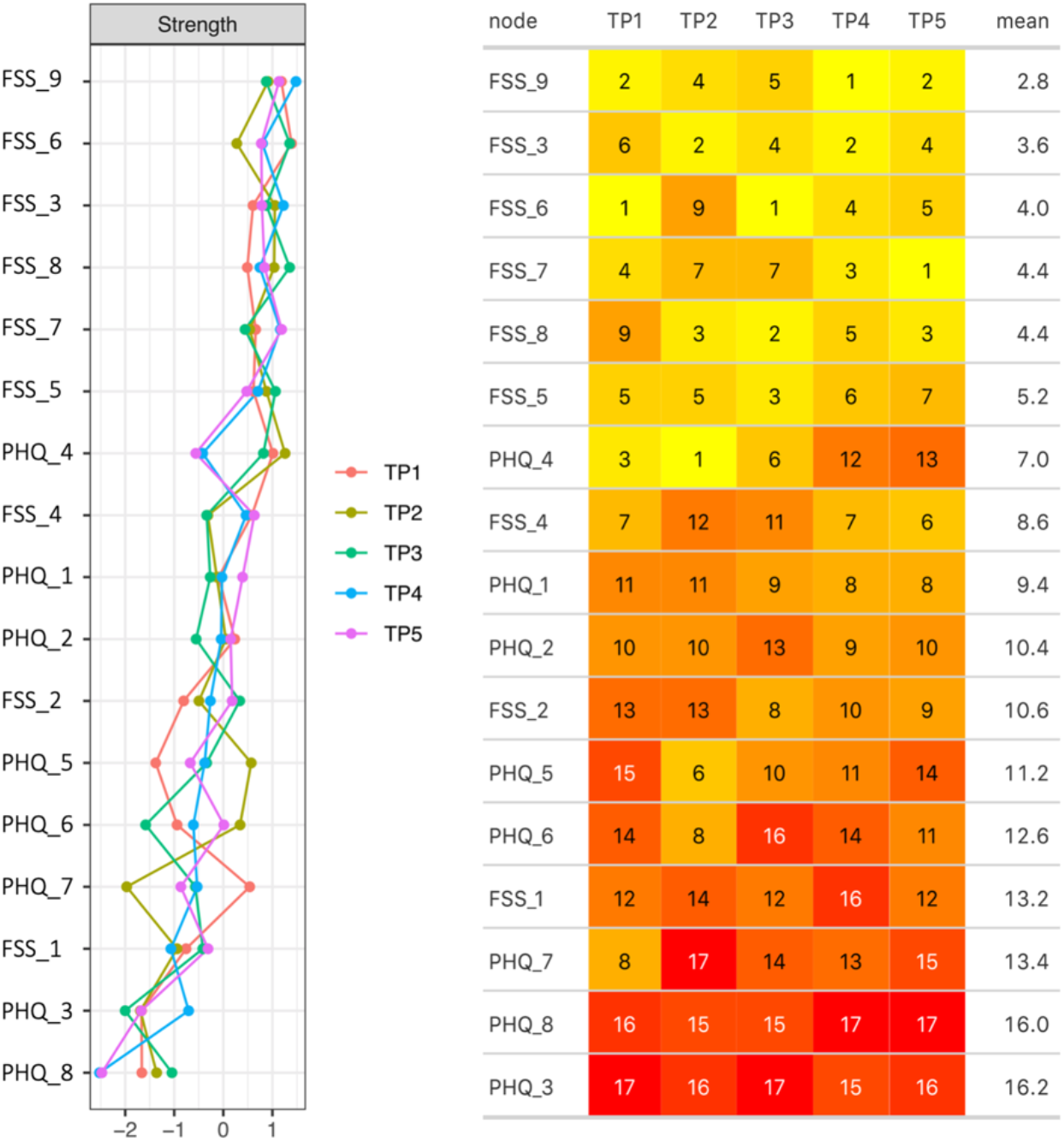
Item strength centrality across time points. Standardized strength node centrality of the 17 FSS/PHQ items across five time points (left), and heatmap table (right) showing the ranked node strength centrality of the five networks, estimated at time point 1-5. Each node is ranked in decreasing order, from 1 (highest centrality), to 17 (lowest centrality). FSS item number 9 (“Fatigue interferes with my work, family, or social life”) demonstrated the highest mean ranked strength centrality across time, followed by FSS item 3 (“I am easily fatigued»).

## Discussion

In a sample of chronic stroke patients, we tested the effect of tDCS combined with computerized cognitive training on fatigue and depressive symptoms. While symptoms of fatigue and depression decreased over the course of the intervention, Bayesian analyses revealed strong evidence of no beneficial effect of tDCS on fatigue or depression severity. To our knowledge, no prior studies have examined *longitudinal* effects of tDCS on PSF in a randomized controlled trial, although a recent study reported reduced FSS scores in stroke patients after a single session of anodal tDCS applied bilaterally to the primary motor cortex (De Doncker et al., 2021). Direct comparison between studies is complicated due to differences in protocols, regarding both electrode montage, stimulation frequency, number of sessions and current amperage. However, there might be greater treatment benefit with higher number of sessions and higher stimulation intensity (Charvet et al., 2018), and we cannot rule out the possibility that we would have observed different results with different frequency and/or intensity. Moreover, tDCS treatment response may interact with individual characteristics such as time since stroke onset, lesion location or lesion size. For example, while Saiote et al. (2014) found no group effects of tDCS on subjective fatigue in patients with MS, a correlation was reported between lesion load in left frontal cortex and treatment response. Future well-powered studies including patients sampled from a wide severity spectrum may be able to discern associations between treatment response and clinical stroke and lesion characteristics.

Non-random attrition is a frequent challenge in clinical intervention studies. Our attrition rate of approximately 26% insinuates that the demands of study participation were unacceptable to a fair proportion of the included patients. On average, the patients who completed the intervention were older and reported lower fatigue scores at baseline compared to the patients who withdrew. We found no group differences in baseline neuropsychological performance, possibly indicating that fatigue constitutes a larger barrier to treatment adherence than cognitive impairments in mildly impaired stroke patients. Importantly, most patients withdrew prior to, and not during the cognitive training, implying that there is no strong basis to infer that the intervention regime was intolerable to patients with fatigue. Rather, one may speculate that patients with high fatigue considered the scope of the intervention in terms of both testing and training to be too demanding, and chose to withdraw at an early stage. Regardless, the observation that more severely fatigued patients withdrew underscores the importance of individually tailoring interventions targeting this patient group. Moreover, the lower age in patients who withdrew from the study may be explained in part by presumably higher family and work obligations among the younger population, conflicting with the time and labor intensity of the intervention.

While the current results provided no evidence of an association between symptoms of fatigue or depression and baseline neuropsychological test performance, both FSS and PHQ were negatively associated with training gain in two and three of six included subtests, respectively. Although effects were small, this mirrors results from our previous findings of no baseline cognitive associations with post stroke fatigue, but evidence of declining performance during a task requiring sustained mental effort (Ulrichsen et al., 2020), corresponding to the concept of cognitive fatiguability (Fiene et al., 2018). While Ulrichsen et al. (2020) examined fatigue effects during a 20 minute attentional response time task, we here extend this finding to the beneficial effects of an intervention spanning several weeks. In contrast to our previous study, the current negative association with training gain was not specific to fatigue, as similar, slightly stronger, association were found for depression. This discrepancy may be due to the difference in performance measures, where the current study targeted change in performance in a range of complex tests over several weeks, reflecting learning rate, while Ulrichsen et al. (2020) measured change in reaction times in a repeated, simple, attentional task at a single session, thus corresponding more closely to the concept of fatiguability (Kluger, Krupp, & Enoka, 2013) or mental fatigue (Johansson & Ronnback, 2014).

The time-resolved network analyses suggested overall higher centrality of fatigue items (except item 1 and 2) than depression items. The central role of fatigue echoes a recent network meta-analysis on depressive symptoms, identifying fatigue as the symptom displaying highest strength centrality across studies (Malgaroli et al., 2021). While the design of the current study does not allow for causal inference, it has been speculated that fatigue exacerbates depression after stroke (Ormstad & Eilertsen, 2015), suggesting that the risk of depression can be reduced with adequate management of fatigue. Following this line of interpretation, the finding that FSS item number 9 – “fatigue interferes with my work, family or social life” displayed the highest strength centrality while simultaneously being among the most stable FSS items across time, may indicate that having fatigue inhibiting social and professional obligations is particularly stressful and predisposes for worsening of respective symptoms. Yet, this hypothesis is based on cross-sectional analyses, and its’ relevance should be tested in future longitudinal studies disentangling the causal relationship between individual symptoms.

Of note, the two FSS items displaying lowest ranked node strength centrality at baseline and across time were item 1 (“my motivation is lower when I am fatigued”) and item 2 (“exercise brings on my fatigue”). This accords with previous reports of poor psychometric properties for item 1 and 2 (Lerdal et al., 2005; Zedlitz, Van, Van, Geurts, & Fasotti, 2016) and improved potential to detect fatigue changes across time after removal of these items (Lerdal & Kottorp, 2011). Our results thus support that these items could be removed in future studies targeting temporal changes in fatigue.

The results should be interpreted considering several limitations. First, the patients suffered from relatively mild strokes, as reflected in the low NIHSS scores, possibly compromising generalizability of results to more severe patient samples. However, both fatigue and depressive complaints were comparable to previous chronic phase stroke studies (Cumming et al., 2018; Dajpratham et al., 2020; Valko, Bassetti, Bloch, Held, & Baumann, 2008), and a substantial portion of the patients reported symptoms above clinical thresholds. Second, as the final assessment of fatigue and depression was collected shortly after the last tDCS stimulation, the current study does not capture potential long-term effects of stimulation, as identified in previous studies (Ayache, Lefaucheur, & Chalah, 2017; Li et al., 2019). Future studies should aim to evaluate long-term effects in addition to immediate response.

Lastly, due to the lack of control group for the cognitive training, we cannot establish whether the observed reduction in fatigue and depressive symptoms was caused by the training, or by other, unknown factors such as anticipation, positive effects of interacting with the research staff, or statistical phenomena such as regression to the mean.

In conclusion, the current study investigated whether tDCS combined with computerized cognitive training alleviated symptoms of fatigue and depression in a sample of chronic phase stroke patients. All though symptoms were reduced during the course of the intervention, Bayesian analysis provided strong evidence for no effect of tDCS compared to sham stimulation. Follow-up analyses of attrition rate, individual differences in training gain and item-level network analyses of fatigue and depression scales support the notion of fatigue as a central clinical symptom, with possible implications for treatment adherence and response.

## Data Availability

The data that support the findings of this study are available from the corresponding author, upon reasonable request.

## Author contributions

**Kristine M. Ulrichsen:** Conceptualization, data collection, formal analyses, figures and tables, writing - original draft and editing, **Knut K. Kolskår:** Experimental design, data collection and curation, writing - review & editing. **Geneviève Richard:** Experimental design, data collection and data curation, writing - review & editing. **Mads Lund Pedersen:** Formal analyses, figures, writing – review & editing. **Dag Alnæs:** Experimental design, writing - review & editing. **Erlend S. Dørum:** Data curation, writing - review & editing. **Anne-Marthe Sanders:** Data collection and data curation, writing - review & editing. **Sveinung Tornås:** Supervision, writing - review & editing. **Luigi Maglanoc:** Formal analyses, interpretation, writing – review & editing. **Andreas Engvig:** interpretation of results, writing - review & editing. **Hege Ihle-Hansen:** interpretation of results, writing - review & editing. **Jan E. Nordvik:** Project administration, funding acquisition, writing - review & editing. **Lars T. Westlye:** Conceptualization, funding acquisition, project administration, supervision, formal writing - review & editing.

## Human Ethics

We obtained appropriate ethical approval from the local ethics committee, and all procedures were in line with the declaration of Helsinki. The study was approved by the Regional Committee for Medical and Health Research Ethics (South-East Norway, 2014/694).

## Acknowledgements

This study was supported by the Norwegian ExtraFoundation for Health and Rehabilitation (<u>2015/FO5146</u>), the Research Council of Norway (<u>249795</u>), the South-Eastern Norway Regional Health Authority (<u>2014097</u>, <u>2015044</u>, <u>2015073</u>, <u>2018037</u>), the European Research Council under the European Union’s Horizon 2020 research and Innovation program (ERC StG Grant 802998), Sunnaas Rehabilitation Hospital and the Department of Psychology, University of Oslo

## References

Ambrus, G. G., Al-Moyed, H., Chaieb, L., Sarp, L., Antal, A., & Paulus, W. (2012). The fade-in–short stimulation–fade out approach to sham tDCS–reliable at 1 mA for naive and experienced subjects, but not investigators. Brain stimulation, 5(4), 499–504.

American Psychiatric Association. (1994). Diagnostic and Statistical Manual of Mental Disorders (4th ed.).

Ayache, S. S., Lefaucheur, J.-P., & Chalah, M. A. (2017). Long term effects of prefrontal tDCS on multiple sclerosis fatigue: A case study. Brain stimulation, 10(5), 1001–1002.

Becker, K. J. (2016). Inflammation and the silent sequelae of stroke. Neurotherapeutics, 13(4), 801–810.

Bringmann, L. F., Elmer, T., Epskamp, S., Krause, R. W., Schoch, D., Wichers, M., … Snippe, E. (2019). What do centrality measures measure in psychological networks? Journal of Abnormal Psychology, 128(8), 892.

Broadbent, D. E., Cooper, P. F., FitzGerald, P., & Parkes, K. R. (1982). The cognitive failures questionnaire (CFQ) and its correlates. British journal of clinical psychology, 21(1), 1–16.

Bucur, M., & Papagno, C. (2018). A systematic review of noninvasive brain stimulation for post-stroke depression. Journal of affective disorders, 238, 69–78.

Bürkner, P.-C. (2017). brms: An R package for Bayesian multilevel models using Stan. Journal of statistical software, 80(1), 1–28.

Charvet, L. E., Dobbs, B., Shaw, M. T., Bikson, M., Datta, A., & Krupp, L. B. (2018). Remotely supervised transcranial direct current stimulation for the treatment of fatigue in multiple sclerosis: results from a randomized, sham-controlled trial. Multiple Sclerosis Journal, 24(13), 1760–1769.

Chen, K., & Marsh, E. B. (2018). Chronic post-stroke fatigue: It may no longer be about the stroke itself. Clinical neurology and neurosurgery, 174, 192–197.

Chen, Y. K., Qu, J. F., Xiao, W. M., Li, W. Y., Weng, H. Y., Li, W., … Ungvari, G. S. (2015). Poststroke fatigue: risk factors and its effect on functional status and health-related quality of life. International journal of stroke, 10(4), 506–512.

Choi-Kwon, S., & Kim, J. S. (2011). Poststroke fatigue: an emerging, critical issue in stroke medicine. International journal of stroke, 6(4), 328–336.

Cumming, T. B., Packer, M., Kramer, S. F., & English, C. (2016). The prevalence of fatigue after stroke: a systematic review and meta-analysis. International journal of stroke, 11(9), 968–977.

Cumming, T. B., Yeo, A. B., Marquez, J., Churilov, L., Annoni, J.-M., Badaru, U., … Lerdal, A. (2018). Investigating post-stroke fatigue: An individual participant data meta-analysis. Journal of psychosomatic research, 113, 107–112.

Dajpratham, P., Pukrittayakamee, P., Atsariyasing, W., Wannarit, K., Boonhong, J., & Pongpirul, K. (2020). The validity and reliability of the PHQ-9 in screening for post-stroke depression. BMC psychiatry, 20(1), 1–8.

De Doncker, W., Dantzer, R., Ormstad, H., & Kuppuswamy, A. (2018). Mechanisms of poststroke fatigue. Journal of Neurology, Neurosurgery & Psychiatry, 89(3), 287–293.

De Doncker, W., Ondobaka, S., & Kuppuswamy, A. (2021). Effect of transcranial direct current stimulation on post-stroke fatigue. Journal of neurology, 1–12.

De Giglio, L., De Luca, F., Prosperini, L., Borriello, G., Bianchi, V., Pantano, P., & Pozzilli, C. (2015). A low-cost cognitive rehabilitation with a commercial video game improves sustained attention and executive functions in multiple sclerosis: a pilot study. Neurorehabilitation and neural repair, 29(5), 453–461.

Delis, D. C. (2000). California verbal learning test. Adult version. Manual. Psychological Corporation.

Delis, D. C., Kaplan, E., & Kramer, J. H. (2001). Delis-Kaplan executive function system.

Douven, E., Köhler, S., Schievink, S.H., van Oostenbrugge, R.J., Staals, J., Verhey, F.R., & Aalten, P. (2017). Temporal associations between fatigue, depression, and apathy after stroke: results of the cognition and affect after stroke, a prospective evaluation of risks study. Cerebrovascular Diseases, 44(5-6), 330-337.

Egorova, N., Cumming, T., Shirbin, C., Veldsman, M., Werden, E., & Brodtmann, A. (2017). Lower cognitive control network connectivity in stroke participants with depressive features. Translational psychiatry, 7(11), 1–8.

Epskamp, S., Borsboom, D., & Fried, E. I. (2018). Estimating psychological networks and their accuracy: A tutorial paper. Behavior Research Methods, 50(1), 195–212.

Epskamp, S., Cramer, A. O. J., Waldorp, L. J., Schmittmann, V. D., & Borsboom, D. (2012). qgraph: Network visualizations of relationships in psychometric data. Journal of statistical software, 48(4), 1–18.

Fiene, M., Rufener, K. S., Kuehne, M., Matzke, M., Heinze, H.-J., & Zaehle, T. (2018). Electrophysiological and behavioral effects of frontal transcranial direct current stimulation on cognitive fatigue in multiple sclerosis. Journal of neurology, 265(3), 607–617.

Filzmoser, P., & Gschwandtner, M. (2018). mvoutlier: Multivariate Outlier Detection Based on Robust Methods. Retrieved from https://CRAN.R-project.org/package=mvoutlier

Fried, E. I., Epskamp, S., Nesse, R. M., Tuerlinckx, F., & Borsboom, D. (2016). What are’good’depression symptoms? Comparing the centrality of DSM and non-DSM symptoms of depression in a network analysis. Journal of affective disorders, 189, 314–320.

Grajny, K., Pyata, H., Spiegel, K., Lacey, E. H., Xing, S., Brophy, C., & Turkeltaub, P. E. (2016). Depression symptoms in chronic left hemisphere stroke are related to dorsolateral prefrontal cortex damage. The Journal of Neuropsychiatry and Clinical Neurosciences, 28(4), 292–298.

Hankey, G. J., Jamrozik, K., Broadhurst, R. J., Forbes, S., & Anderson, C. S. (2002). Long-term disability after first-ever stroke and related prognostic factors in the Perth Community Stroke Study, 1989–1990. Stroke, 33(4), 1034–1040.

Johansson, B., & Ronnback, L. (2014). Evaluation of the mental fatigue scale and its relation to cognitive and emotional functioning after traumatic brain injury or stroke. Int J Phys Med Rehabil, 2(01).

Kjeverud, A., Østlie, K., Schanke, A.-K., Gay, C., Thoresen, M., & Lerdal, A. (2020). Trajectories of fatigue among stroke patients from the acute phase to 18 months post-injury: A latent class analysis. Plos one, 15(4), e0231709.

Kluger, B. M., Krupp, L. B., & Enoka, R. M. (2013). Fatigue and fatigability in neurologic illnesses: proposal for a unified taxonomy. Neurology, 80(4), 409–416.

Kolskaar, K. K., Richard, G., Alnaes, D., Dørum, E. S., Sanders, A. M., Ulrichsen, K. M., … Westlye, L. T. (2020). Reliability, sensitivity, and predictive value of fMRI during multiple object tracking as a marker of cognitive training gain in combination with tDCS in stroke survivors. Human Brain Mapping.

Kroenke, K., Spitzer, R. L., & Williams, J. B. W. (2001). The PHQ-9: validity of a brief depression severity measure. Journal of general internal medicine, 16(9), 606–613.

Krupp, L. B., LaRocca, N. G., Muir-Nash, J., & Steinberg, A. D. (1989). The fatigue severity scale: application to patients with multiple sclerosis and systemic lupus erythematosus. Archives of neurology, 46(10), 1121–1123.

Leegaard, O. F. (1983). Diffuse cerebral symptoms in convalescents from cerebral infarction and myocardial infarction. Acta Neurologica Scandinavica, 67(6), 348–355.

Lerdal, A., & Kottorp, A. (2011). Psychometric properties of the Fatigue Severity Scale— Rasch analyses of individual responses in a Norwegian stroke cohort. International journal of nursing studies, 48(10), 1258–1265.

Lerdal, A., Wahl, A. K., Rustoen, T., Hanestad, B. R., & Moum, T. (2005). Fatigue in the general population: a translation and test of the psychometric properties of the Norwegian version of the fatigue severity scale. Scandinavian journal of public health, 33(2), 123–130.

Li, M.-S., Du, X.-D., Chu, H.-C., Liao, Y.-Y., Pan, W., Li, Z., & Hung, G. C.-L. (2019). Delayed effect of bifrontal transcranial direct current stimulation in patients with treatment-resistant depression: a pilot study. BMC psychiatry, 19(1), 1–9.

Llorca, G. E., Castilla-Guerra, L., Moreno, M. C. F., Doblado, S. R., & Hernández, M. D. J. (2015). Post-stroke depression: an update. Neurología (English Edition), 30(1), 23–31.

Malgaroli, M., Calderon, A., & Bonanno, G. A. (2021). Networks of major depressive disorder: A systematic review. Clinical Psychology Review, 102000.

Meyer, B. C., & Lyden, P. D. (2009). The modified National Institutes of Health Stroke Scale: its time has come. International journal of stroke, 4(4), 267–273.

Michael, K. (2002). Fatigue and stroke. Rehabilitation nursing, 27(3), 89–94.

Morsund, Å.H., Ellekjær, H., Gramstad, A., Reiestad, M. T., Midgard, R., Sando, S. B., … Næss, H. (2019). The development of cognitive and emotional impairment after a minor stroke: A longitudinal study. Acta Neurologica Scandinavica, 140(4), 281–289.

Maaijwee, N. A., Arntz, R. M., Rutten-Jacobs, L. C., Schaapsmeerders, P., Schoonderwaldt, H. C., van Dijk, E. J., & de Leeuw, F.-E. (2015). Post-stroke fatigue and its association with poor functional outcome after stroke in young adults. Journal of Neurology, Neurosurgery & Psychiatry, 86(10), 1120–1126.

Nadarajah, M., & Goh, H.-T. (2015). Post-stroke fatigue: a review on prevalence, correlates, measurement, and management. Topics in stroke rehabilitation, 22(3), 208–220.

Naess, H., Lunde, L., Brogger, J., & Waje-Andreassen, U. (2012). Fatigue among stroke patients on long-term follow-up. The Bergen Stroke Study. Journal of the neurological sciences, 312(1-2), 138–141.

Nasreddine, Z. S., Phillips, N. A., Bédirian, V., Charbonneau, S., Whitehead, V., Collin, I., … Chertkow, H. (2005). The Montreal Cognitive Assessment, MoCA: a brief screening tool for mild cognitive impairment. Journal of the American Geriatrics Society, 53(4), 695–699.

Nguyen, S., Wong, D., McKay, A., Rajaratnam, S. M. W., Spitz, G., Williams, G., … Ponsford, J. L. (2019). Cognitive behavioural therapy for post-stroke fatigue and sleep disturbance: a pilot randomised controlled trial with blind assessment. Neuropsychological rehabilitation, 29(5), 723–738.

Ormstad, H., & Eilertsen, G. (2015). A biopsychosocial model of fatigue and depression following stroke. Medical hypotheses, 85(6), 835–841.

Passier, P., Post, M., van Zandvoort, M., Rinkel, G., Lindeman, E., & Visser-Meily, J. (2011). Predicting fatigue 1 year after aneurysmal subarachnoid hemorrhage. Journal of neurology, 258(6), 1091–1097.

Pérez-Martín, M. Y., González-Platas, M., Eguía-del Río, P., Croissier-Elías, C., & Sosa, A. J. (2017). Efficacy of a short cognitive training program in patients with multiple sclerosis. Neuropsychiatric Disease and Treatment, 13, 245.

Pihlaja, R., Uimonen, J., Mustanoja, S., Tatlisumak, T., & Poutiainen, E. (2014). Post-stroke fatigue is associated with impaired processing speed and memory functions in first-ever stroke patients. Journal of psychosomatic research, 77(5), 380–384.

Ponchel, A., Bombois, S., Bordet, R., & Hénon, H. (2015). Factors associated with poststroke fatigue: a systematic review. Stroke research and treatment, 2015.

R core team. (2020). R: A language and environment for statistical computing. R Foundation for Statistical Computing, Vienna, Austria. Retrieved from https://www.R-project.org/.

Radman, N., Staub, F., Aboulafia-Brakha, T., Berney, A., Bogousslavsky, J., & Annoni, J.-M. (2012). Poststroke fatigue following minor infarcts: a prospective study. Neurology, 79(14), 1422–1427.

Richard, G., Petersen, A., Ulrichsen, K. M., Kolskår, K. K., Alnæs, D., Sanders, A.-M., … Westlye, L. T. (2020). TVA-based modeling of short-term memory capacity, speed of processing and perceptual threshold in chronic stroke patients undergoing cognitive training: case-control differences, reliability, and associations with cognitive performance. PeerJ, 8, e9948.

Saiote, C., Goldschmidt, T., Timäus, C., Steenwijk, M. D., Opitz, A., Antal, A., … Nitsche, M. A. (2014). Impact of transcranial direct current stimulation on fatigue in multiple sclerosis. Restorative neurology and neuroscience, 32(3), 423–436.

Schepers, V. P., Visser-Meily, A. M., Ketelaar, M., & Lindeman, E. (2006). Poststroke fatigue: course and its relation to personal and stroke-related factors. Archives of physical medicine and rehabilitation, 87(2), 184–188.

Snaphaan, L., Van der Werf, S., & de Leeuw, F. E. (2011). Time course and risk factors of post-stroke fatigue: a prospective cohort study. European Journal of Neurology, 18(4), 611–617.

Starkstein, S. E., & Hayhow, B. D. (2019). Treatment of post-stroke depression. Current treatment options in neurology, 21(7), 1–10.

Tang, W. K., Chen, Y. K., Liang, H. J., Chu, W. C. W., Mok, V. C. T., Ungvari, G. S., & Wong, K. S. (2014). Subcortical white matter infarcts predict 1-year outcome of fatigue in stroke. BMC neurology, 14(1), 1–6.

Tang, W. K., Chen, Y. K., Mok, V., Chu, W. C., Ungvari, G. S., Ahuja, A. T., & Wong, K. S. (2010). Acute basal ganglia infarcts in poststroke fatigue: an MRI study. Journal of neurology, 257(2), 178–182.

Ulrichsen, K. M., Alnæs, D., Kolskår, K. K., Richard, G., Sanders, A. M., Dørum, E. S., … Nordvik, J. E. (2020). Dissecting the cognitive phenotype of post-stroke fatigue using computerized assessment and computational modeling of sustained attention. European Journal of Neuroscience.

Valiengo, L., Casati, R., Bolognini, N., Lotufo, P. A., Benseñor, I. M., Goulart, A. C., & Brunoni, A. R. (2016). Transcranial direct current stimulation for the treatment of post-stroke depression in aphasic patients: a case series. Neurocase, 22(2), 225–228.

Valiengo, L. C. L., Goulart, A. C., de Oliveira, J. F., Benseñor, I. M., Lotufo, P. A., & Brunoni, A. R. (2017). Transcranial direct current stimulation for the treatment of post-stroke depression: results from a randomised, sham-controlled, double-blinded trial. Journal of Neurology, Neurosurgery & Psychiatry, 88(2), 170–175.

Valko, P. O., Bassetti, C. L., Bloch, K. E., Held, U., & Baumann, C. R. (2008). Validation of the fatigue severity scale in a Swiss cohort. Sleep, 31(11), 1601–1607.

van de Port, I. G., Kwakkel, G., Bruin, M., & Lindeman, E. (2007). Determinants of depression in chronic stroke: a prospective cohort study. Disability and rehabilitation, 29(5), 353–358.

Walsh, M. E., Galvin, R., Loughnane, C., Macey, C., & Horgan, N. F. (2015). Community re-integration and long-term need in the first five years after stroke: results from a national survey. Disability and rehabilitation, 37(20), 1834–1838.

Wang, S.-B., Wang, Y.-Y., Zhang, Q.-E., Wu, S.-L., Ng, C. H., Ungvari, G. S., … Xiang, Y.-T. (2018). Cognitive behavioral therapy for post-stroke depression: a meta-analysis. Journal of affective disorders, 235, 589–596.

Wechsler. (2011). T. N. P. San Antonio.

Whitehead, L. (2009). The measurement of fatigue in chronic illness: a systematic review of unidimensional and multidimensional fatigue measures. Journal of pain and symptom management, 37(1), 107-128.

Wu, S., Kutlubaev, M. A., Chun, H. Y. Y., Cowey, E., Pollock, A., Macleod, M. R., … Mead, G. E. (2015). Interventions for post-stroke fatigue. Cochrane Database of Systematic Reviews(7).

Wu, S., Mead, G., Macleod, M., & Chalder, T. (2015). Model of understanding fatigue after stroke. stroke, 46(3), 893–898.

Zedlitz, A., Van, M. M., Van, E. M., Geurts, A., & Fasotti, L. (2016). Psychometric properties of FSS and CIS-20r for measuring post-stroke fatigue. International Journal of Psychology, 2, 27.

Aarnes, R., Stubberud, J., & Lerdal, A. (2020). A literature review of factors associated with fatigue after stroke and a proposal for a framework for clinical utility. Neuropsychological rehabilitation, 30(8), 1449–1476.

