## Supplementary Material for "No effect of tDCS on fatigue and depression in chronic stroke patients: an exploratory randomized sham-controlled trial combining tDCS with computerized cognitive training"

**Supplementary Figure 1.** Baseline FSS and PHQ scores plotted against Cogmed slopes

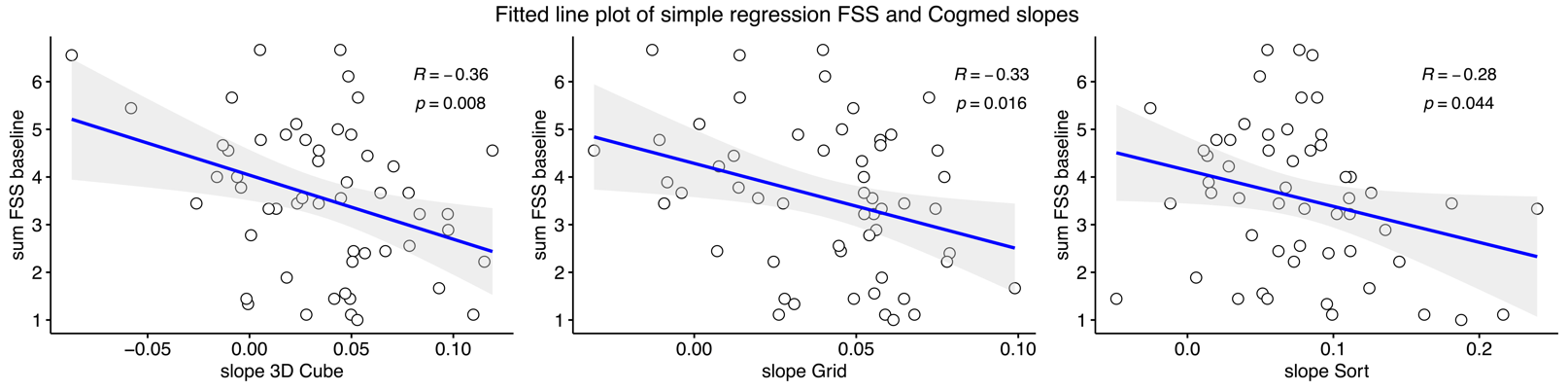

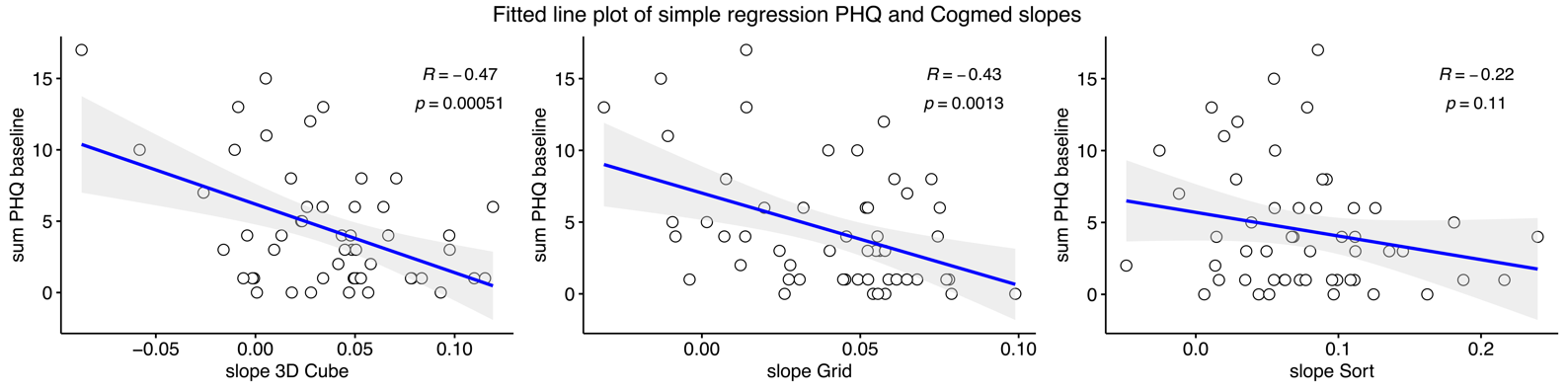

**Supplementary Table 1.** Mean aggregated scores (TP1 – TP5) and CV values for FSS and PHQ items.

|  | *Fatigue Severity Scale* |  |  |  |
| --- | --- | --- | --- | --- |
|  | Score range 1 - 7 | Mean | SD | CV |
| 1 | My motivation is lower when I am fatigued | 22.5 | 7.4 | 31 |
| 2 | Exercise brings on my fatigue | 14.3 | 6.9 | 39 |
| 3 | I am easily fatigued | 15.4 | 6.9 | 36 |
| 4 | Fatigue interferes with my physical functioning | 19.5 | 6.7 | 33 |
| 5 | Fatigue causes frequent problems for me | 13.1 | 6.0 | 37 |
| 6 | My fatigue prevents sustained physical functioning | 15.7 | 7.3 | 36 |
| 7 | Fatigue interferes with carrying out certain duties and responsibilities | 16.7 | 7.0 | 35 |
| 8 | Fatigue is among my most disabling symptoms | 16.3 | 8.0 | 34 |
| 9 | Fatigue interferes with my work, family, or social life | 15.1 | 7.0 | 32 |
|  | *Patient Health Questionnaire (PHQ – 9)* |  |  |  |
|  | Score range 0 -3 | Mean | SD | CV |
| 1 | Little interest or pleasure in doing things | 2.4 | 2.6 | 20 |
| 2 | Feeling down, depressed, or hopeless | 6.3 | 5.6 | 16 |
| 3 | Trouble falling or staying asleep, or sleeping too much | 8.0 | 6.3 | 23 |
| 4 | Feeling tired or having little energy | 7.8 | 6.2 | 22 |
| 5 | Poor appetite or overeating | 5.2 | 5.4 | 14 |
| 6 | Feeling bad about yourself — or that you are a failure or have let yourself or your family down | 2.1 | 2.3 | 19 |
| 7 | Trouble concentrating on things, such as reading the newspaper or watching television | 1.9 | 2.4 | 17 |
| 8 | Moving or speaking so slowly that other people could have noticed? Or the opposite — being so fidgety or restless that you have been moving around a lot more than usual | 1.0 | 1.5 | 12 |
| 9 | Thoughts that you would be better off dead or of hurting yourself in some way | 0.3 | 1.0 | 4 |

*CV = coefficient of variation. We estimated one across-time CV value per item for each patient, and CV value in the table is the *mean* of these individual CV values.

**Supplementary Figure 2.** Bootstrapped difference test for edges (left) and node strength centrality (right), for basline network esimtated with FSS sum score and all PHQ items.

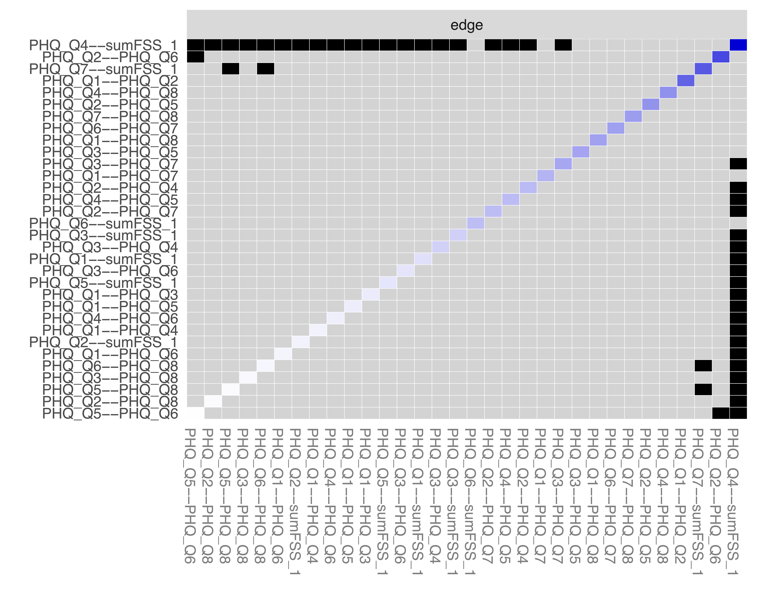

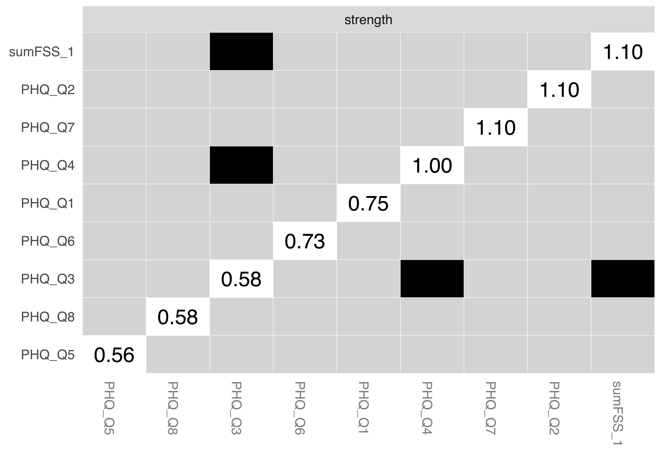

**Supplementary Figure 3.** Bootstrapped difference test for edges in full (all-item) baseline network.

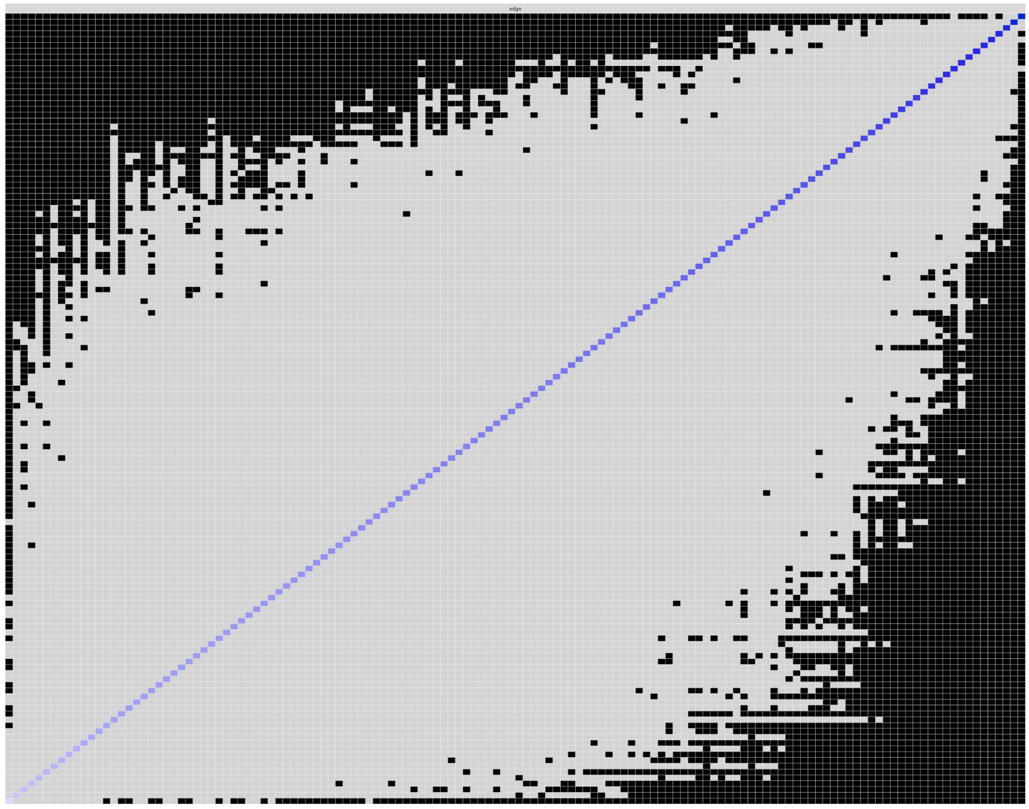

Bootstrapped edge difference tests (*α* = 0.05) between non-zero edge-weights (153 in total) in the estimated network from baseline. Names have been removed due to abundancy. Grey boxes indicate that edges do not differ significantly from other edges.

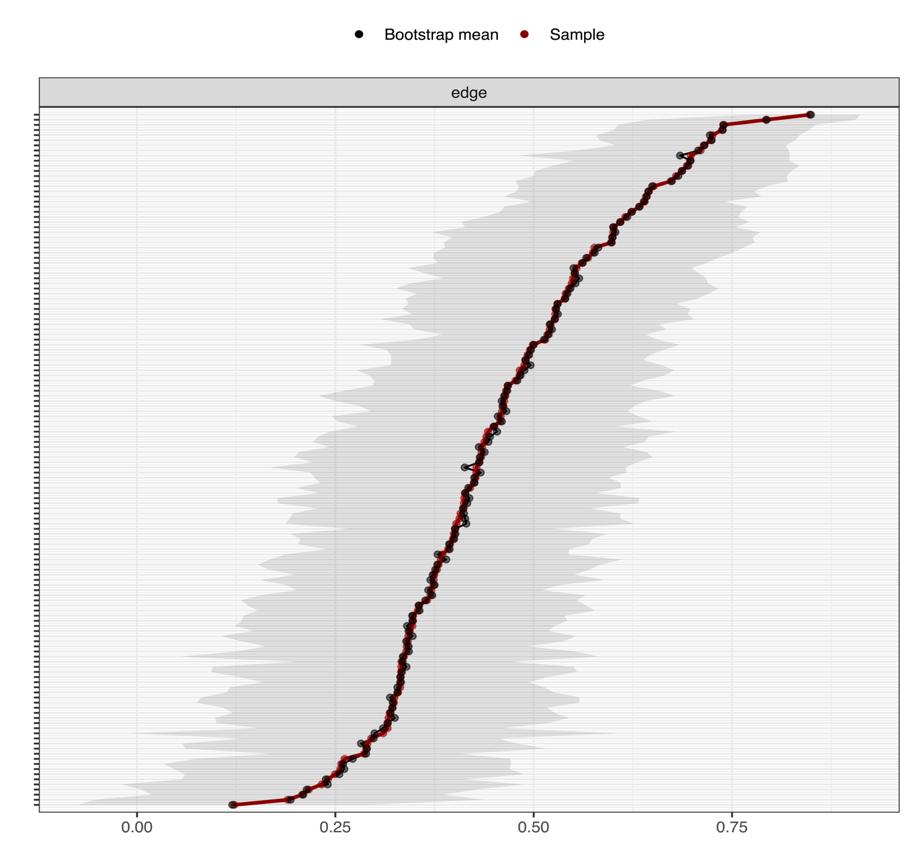
**Supplementary Figure 4.** Bootstrapped CIs around edge-weights

**Figure 4.** Bootstrapped CIs (grey area) around estimated edge-weights in the baseline network. Each line on the y-axis represent an edge (names have been removed due to abundancy). Most CIs are of considerable size and overlap with other edge-weights and suggests that interpretation of edge order should be done with caution.

**Supplementary Figure 5.** Unregularized, individual item networks of FSS and PHQ scores for each time point. Node placement is defined by loadings on (unrotated) PCA dimensions.

TP1	TP2

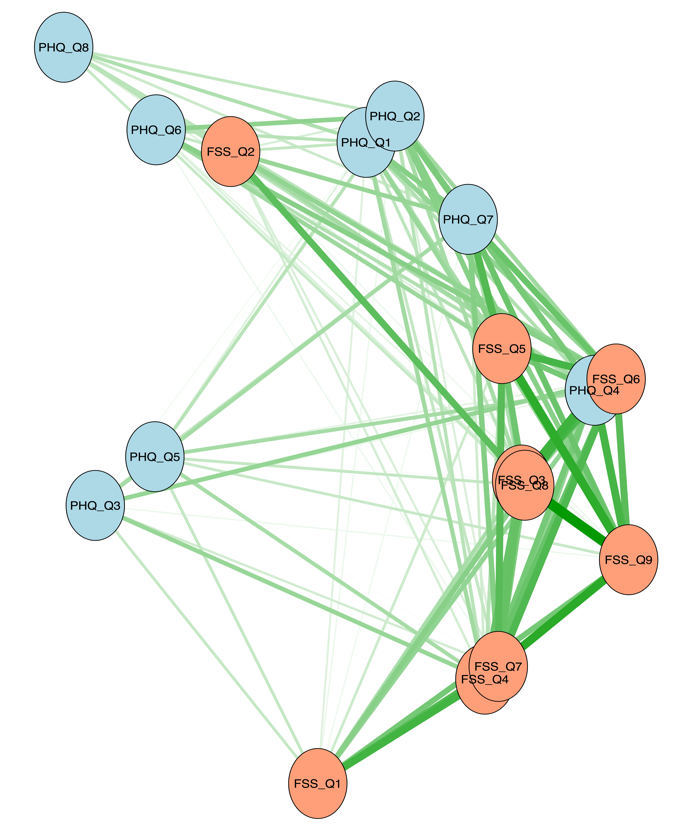

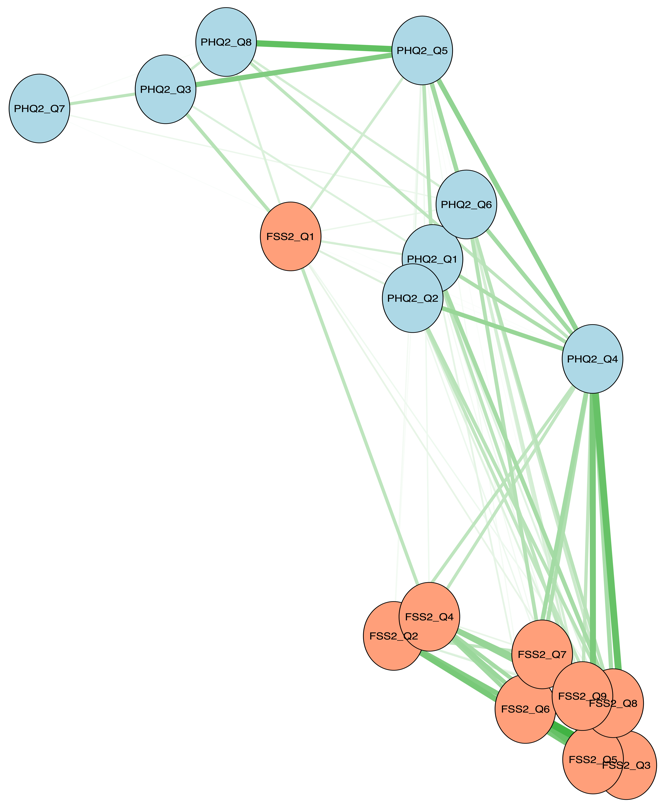

TP3	TP4

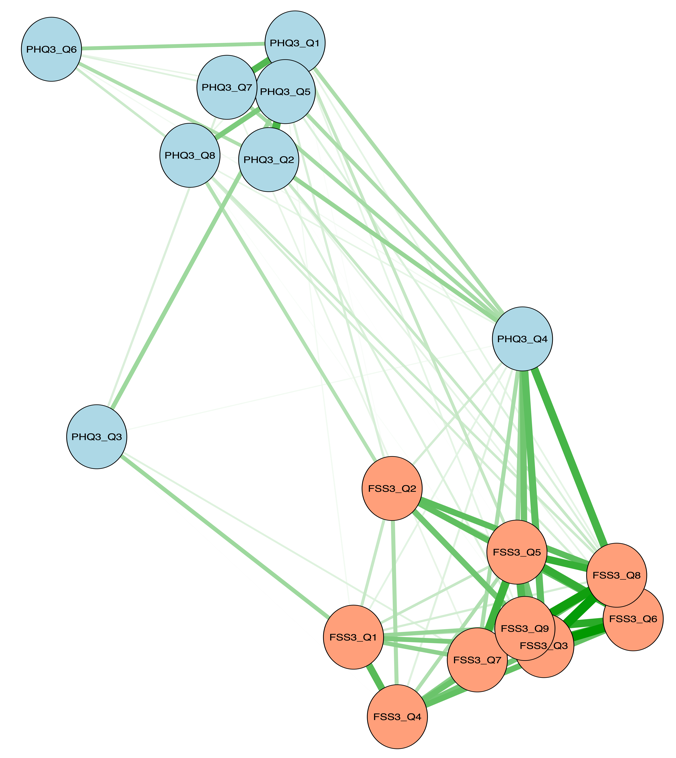

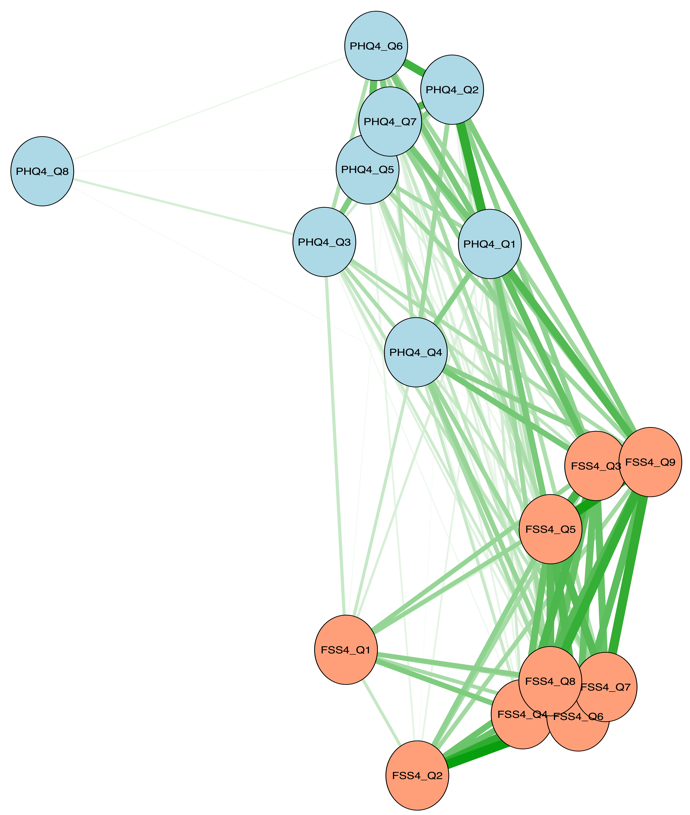

TP5

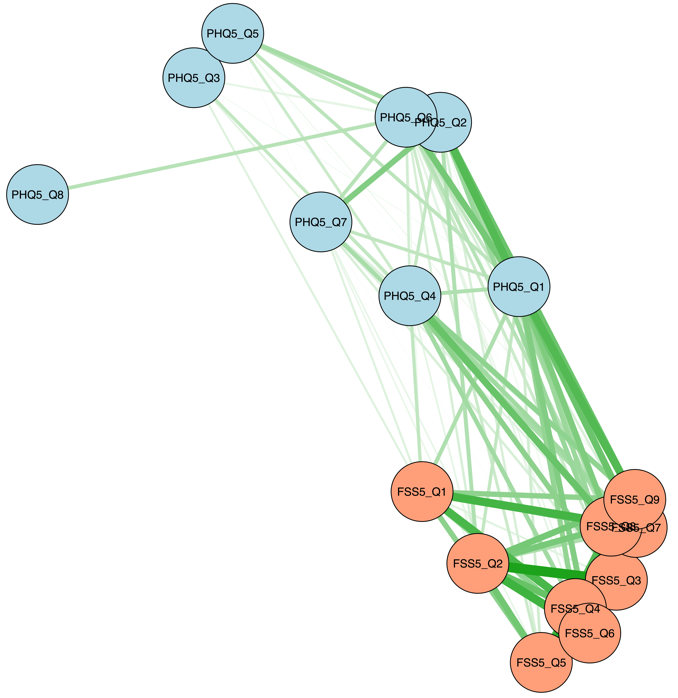

**Supplementary Figure 6.** Bootstrapped difference tests for node strength centrality in all-item networks, time-point 1 to 5.

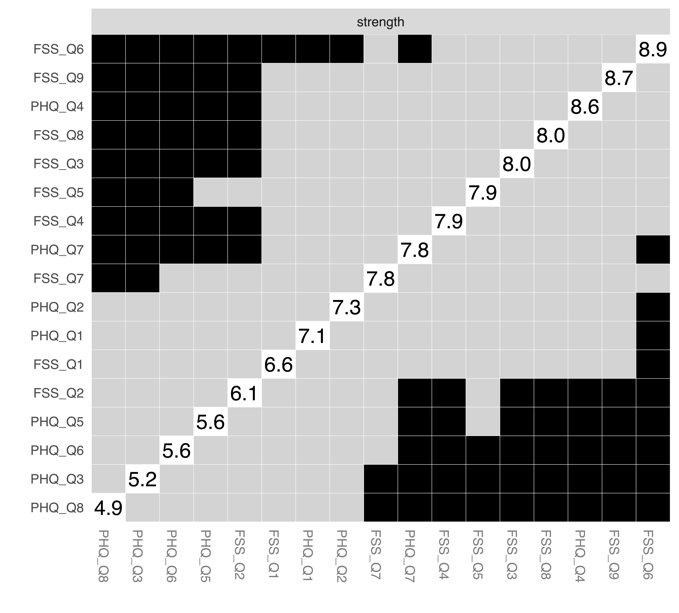

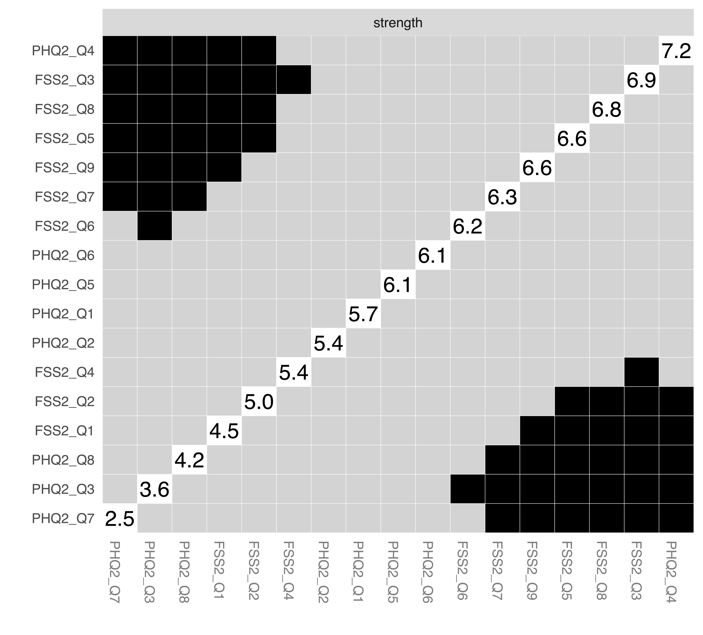
 TP1	TP2

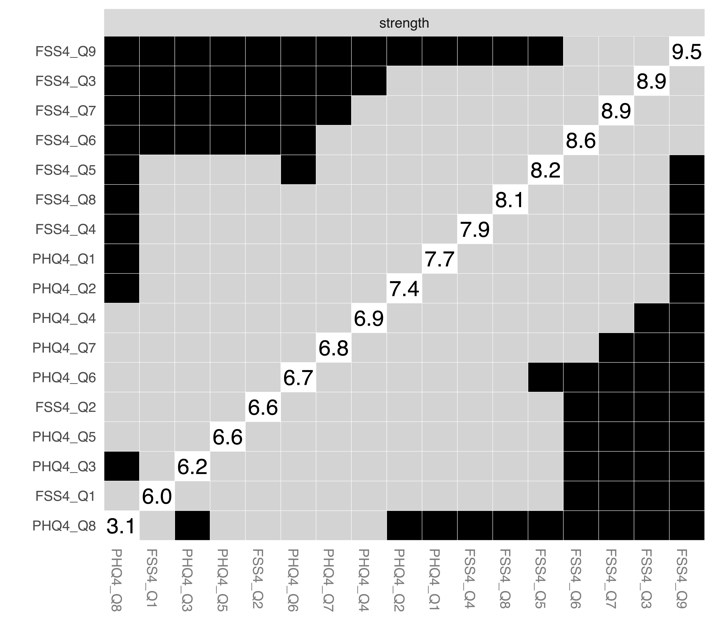

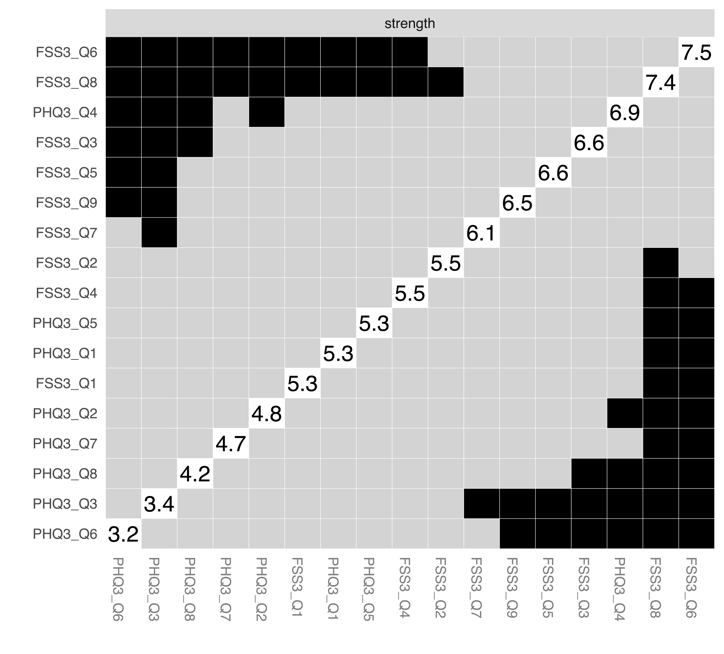
 TP3	TP4

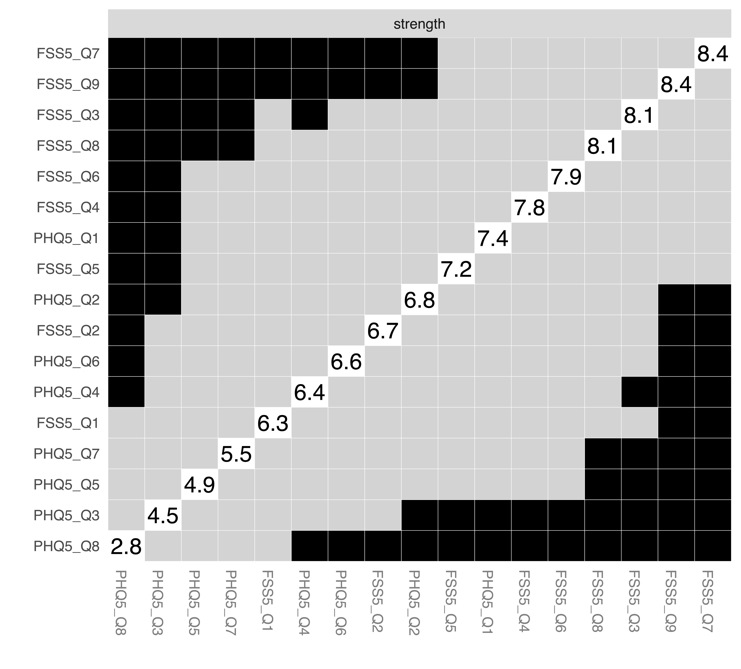
 TP5

| **Table 2.** | Bayes factor estimations for linear correlations (default priors) | | | |
| --- | --- | --- | --- | --- |
| FSS |  | **Mean posterior*** | **BF**_10_****** | **97.5 % CI** |
|  | WASI | 0.10 | 0.42 | -0.13 – 0.34 |
|  | MoCA | -0.02 | 0.27 | -0.24 – 0.22 |
|  | CVLT total recall | 0.02 | 0.27 | -0.20 – 0.24 |
|  | Stroop inhibiton | -0.00 | 0.28 | -0.23 – 0.22 |
|  | Stroop switch | -0.07 | 0.35 | -0.30 – 0.15 |
|  | CFQ | 0.41 | >100 | 0.21 – 0.59 |
| PHQ |  | **Mean posterior** | **BF**_10_ | **97.5 % CI** |
|  | WASI | 0.10 | 0.42 | -0.12 – 0.33 |
|  | MoCA | 0.10 | 0.41 | -0.12 – 0.33 |
|  | CVLT total recall | 0.01 | 0.27 | -0.22 – 0.24 |
|  | Stroop inhibiton | 0.04 | 0.30 | -0.19 – 0.27 |
|  | Stroop switch | -0.04 | 0.30 | -0.28 – 0.19 |
|  | CFQ | 0.52 | >100 | 0.32 – 0.68 |

** BF_10_ is the Bayes factor giving the evidence for *H*1 over *H*0. BF_10_ < 1 provides no evidence for *H*1 over *H*0, while BF_10_ > 100 indicates very strong evidence for *H*1 over *H*0.	* Mean posterior corresponds to correlation coefficient

B:

Represents the correlation of the centrality of nodes in the original network (Figure 1A) with the centrality of net-

works sampled while dropping participants. When the correlation after dropping a substantial amount of participants

is high, it means the centrality estimates in the original network can be considered stable.
